# A global omics data sharing and analytics marketplace: Case study of a rapid data COVID-19 pandemic response platform

**DOI:** 10.1101/2020.09.28.20203257

**Authors:** Ibrahim Farah, Giada Lalli, Darrol Baker, Axel Schumacher

**Affiliations:** Shivom Ventures Limited, 71-75 Shelton Street, Covent Garden, London, England, WC2H 9JQ; Division of Psychiatry, University College London; Katholieke Universiteit Leuven, Belgium; GoldenHelix Foundation, London. England, WC2N 5AP; Novo Nordisk Research Centre Oxford, Innovation Building, Old Road Campus, Roosevelt Drive, Oxford, UK

**Keywords:** Data sharing, Cohort studies, COVID-19, Omics data, Genomics, Pandemic preparedness, SARS-CoV2, Bioinformatics, Data marketplace, Precision medicine

## Abstract

Under public health emergencies, particularly an early epidemic, it is fundamental that genetic and other healthcare data is shared across borders in both a timely and accurate manner before the outbreak of a global pandemic. However, although the COVID-19 pandemic has created a tidal wave of data, most patient data is siloed, not easily accessible, and due to low sample size, largely not actionable. Based on the precision medicine platform Shivom, a novel and secure data sharing and data analytics marketplace, we developed a versatile pandemic preparedness platform that allows healthcare professionals to rapidly share and analyze genetic data. The platform solves several problems of the global medical and research community, such as siloed data, cross-border data sharing, lack of state-of-the-art analytic tools, GDPR-compliance, and ease-of-use. The platform serves as a central marketplace of ‘discoverability’. The platform combines patient genomic & omics data sets, a marketplace for AI & bioinformatics algorithms, new diagnostic tools, and data-sharing capabilities to advance virus epidemiology and biomarker discovery. The bioinformatics marketplace contains some preinstalled COVID-19 pipelines to analyze virus- and host genomes without the need for bioinformatics expertise. The platform will be the quickest way to rapidly gain insight into the association between virus-host interactions and COVID-19 in various populations which can have a significant impact on managing the current pandemic and potential future disease outbreaks.

## Introduction

In December 2019, a cluster of severe pneumonia cases epidemiologically linked to an open-air live animal market in the city of Wuhan, China ^1,2^. The sudden spread of this deadly disease, later called COVID-19, led local health officials to issue an epidemiological alert to the World Health Organization’s (WHO) and the Chinese Center for Disease Control and Prevention. Quickly, the etiological agent of the unknown disease was found to be a coronavirus, SARS-CoV-2, the same subgenus as the SARS virus that caused a deadly global outbreak in 2003. By reconstructing the evolutionary history of SARS-CoV-2, an international research team has discovered that the lineage that gave rise to the virus has been circulating in horseshoe bats for decades and likely includes other viruses with the ability to infect humans^3^. The team concluded that the global health system was too late in responding to the initial SARS-CoV-2 outbreak, and that preventing future pandemics will require the implementation of human disease surveillance systems that are able to quickly identify novel pathogens and their interaction with the humans host and respond in real time. The key to successful surveillance is knowing which viruses caused the new outbreak and how the host genome affects severity of disease. In outbreaks of zoonotic pathogens, identification of the infection source is crucial because this may allow health authorities to separate human populations from the pathogen source (e.g. wildlife or domestic animal reservoirs) posing the zoonotic risk^3^. Even if stopping an outbreak in its early stages is not possible, data collection, data sharing, and analysis of host<>virus interactions is nevertheless important for containment purposes in other regions of the planet and prevention of future outbreaks. Such surveillance measures are easily implemented with the pandemic preparedness platform showcased in this report that is based on the novel data sharing and data analytics marketplace <u>Shivom</u>^4^ (https://www.shivom.iso/). The platform is a proven research ecosystem used by universities, biotech, and bioinformatics organizations to share and analyze omics data and can be used for a variety of use cases; from precision medicine, drug discovery, translational science to building data repositories, and tackling a disease outbreak.

In the current coronavirus outbreak, the global research community is mounting a large-scale public health response to understand the pathogen and combat the pandemic. Vast amounts of new data are being generated and need to be analyzed in the context of pre-existing data and shared for solution-centered research approaches to combat the disease and inform public health policies. However, research showed that the vast majority of clinical studies on COVID-19 do not meet proper quality standards due to small sample size^5^.

Our approach is designed to provide healthcare professionals with an urgently needed platform to find and analyze genetic data, and securely and anonymously share sensitive patient data to fight the disease outbreak. No patient data can be traced back to the data donor (patient or data custodian) because only algorithms can access the raw data. Researchers querying the data-hub have no access to the genetic data source, i.e. no raw data (e.g. no sequencing reads can be downloaded). Instead, researchers and data analysts can run predefined bioinformatics and AI algorithms on the data. Only summary statistics are provided (e.g. GWAS, PHEWAS, polygenic risk score analyses). In addition to genetics bioinformatics pipelines, specialized COVID-19 bioinformatics tools are available (e.g., assembly, mapping, and metagenomics pipelines). Using these OPEN tools, researchers can quickly understand why some people develop severe illness while others do show only mild symptoms or no symptoms whatsoever. We might hypothesize that some people harbor protective gene variants, or their gene regulation reacts differently when the COVID-19 virus attacks the host.

### Breaking down data silos

The usability of digital healthcare data is limited when it is trapped in silos limiting its maximum potential value. Examples of COVID-19 data depositories include the European Commission COVID-19 Data Portal^6^, the Canadian COVID Genomics Network^7^ (CanCOGeN), Genomics England and the GenOMICC (Genetics of Mortality in Critical Care) consortium^8^, the COVID-19 Host Genetics Initiative^9^, the National COVID Cohort Collaborative (N3C)^10^, the COVID-19 Genomics UK (COG-UK) Consortium^11^, the Secure Collective COVID-19 Research (SCOR) consortium^12^, the International COVID-19 Data Research Alliance^13^, and the Covid Human Genetic Effort consortium^14^, among several others. All of them, while very valuable for fighting the pandemic, present complicated access hurdles, limited data-sharing capabilities, and missing interoperability and analytics capabilities that limit their usefulness.

Compartmentalized information limits our healthcare system’s ability to make large strides in research and development restricting the available benefits that can be generated for the public. This is especially true for omics data. If omics data silos are broken up and can be globally aggregated, we believe that society will realize a gain of value that observes accelerating returns. If individuals and medical centers around the world shared their genomic data more fully, then as a whole this data would be significantly more useful. Particularly the role of host genetics in impacting susceptibility and severity of COVID-19 has been studied only in a few small cohorts. Expanding the opportunities to find patients somewhere in the world that possess unusual mutations and phenotypes relevant to national efforts, or have SARS-CoV-2 resistant mutations, would shift precision medicine to another level.

By combining genome sequenced data with health records, researchers and clinicians would have an invaluable resource that can be used to improve patient outcomes, and additionally be used to investigate the causes and treatments of diseases. For big data approaches to thrive, however, historical barriers dividing research groups and institutions need to be broken down and a new era of open, collaborative and data-driven science needs to be ushered in.

There is also a cost to using and producing data, and if this cost is shared amongst many stakeholder’s new data sources, which outperform older databases significantly, can be built. Another obstacle is data generation itself. Genomes have to come from somewhere and currently researchers have limited access to them. Healthcare organizations and private individuals often have restricted access to their own data and even when they do, they are limited in the ways they can use this data, or for sharing with third parties (e.g. for scientific studies). Computer scientists, bioinformaticians, and machine learning researchers are tackling the pandemic the way they know how: compiling datasets and building algorithms to learn from them. But the vast majority of data collected is not accessible to the public, partly because there is no platform that offers anonymized data sharing on a global level, while addressing privacy concerns, and regulatory hurdles. What is needed is a platform that can store data in a secure, anonymized, and impenetrable manner with the goal of preventing malicious attacks or unauthorized access. While ensuring access controls are maintained and satisfied, the database must also be user friendly, publicly accessible, and searchable.

In addition, there must be ways to quickly and easily analyze the collected data. These properties make the data in the database usable, meaning it must be easy for use in population health studies, pharmaceutical R&D, and personal genomics. In addition, such a platform must be accessible on a global scale, as well as offer data provenance and auditing features. We believe that for such a platform to be successful, adding new genomic data to the platform must be trivial and accessible to everyone. We aim to provide such a platform by using convergent technologies including genomics, cryptography, distributed ledgers, and artificial intelligence.

### Data sharing during a pandemic

Global pandemics come and go. The tragic impacts of the unusually deadly influenza pandemic in 1918, called the Spanish flu, or the bubonic plague that killed as many as one-third of Europe’s and North Africa’s population in the 14^th^ century reminds us that COVID-19 wasn’t the first global pandemic, and it is clear that it won’t be the last. In contrast to the Middle Ages, we now have the technological tools and exceptional scientific networks to fight pandemics. Already, the COVID-19 pandemic has demonstrated that sharing data can improve clinical outcomes for patients, and that quick access to data literally has life-or-death consequences in healthcare delivery. However, only a minority of healthcare organizations are participating in data-sharing efforts. Without proper coordination amongst initiatives and agencies, and making the efforts public, these initiatives run the risk of duplicating efforts or missing opportunities, resulting in slower progress and economic inefficiencies and further data silos.

Comprehensive solutions to problems such as the SARS-CoV-2 virus outbreak require genuine international cooperation, not only between governments and the private sector, but also from all levels of our health systems. Researchers from all over the world need to be able to act quickly, without waiting for international efforts to slowly coordinate their efforts, to understand early local outbreaks and to start intervention measures without delay. New solutions to tackle the pandemic need also early adopters that want to move away from our archaic data information systems that can hamstring real-time detection of emerging disease threats.

### The need for rapid open data sharing

Many research and innovation actors have reoriented some of their previously funded activities towards COVID-19, but often with little guidance from policy makers. Despite all complications and concerns, the advantages of sharing data quickly and safely can be significant. A lack of coordination at international level on numerous initiatives launched by different institutions to combat COVID-19 resulted in notably reduced cooperation and exchanges of data and results between projects, limited interoperability, as well as lower data quality and interpretation. Open data plays a major role in the global response to large-scale outbreaks. For instance, analysis of the 2016 Ebola outbreak points to the importance of open data, including genomic data to generate learnings about diseases where effective vaccines are lacking or have not been developed. Similarly, open data played a major role in the response to the 2016 Zika outbreak with a commitment from leading national agencies and science organizations to share data. With the Shivom platform, it is possible to help the global research community fast-track discover new preventive measures and optimize logistic solutions. Moreover, this data lends itself naturally in predicting and preparing for future, inevitable disease outbreaks.

A commitment to open access in genomics research has found widespread backing in science and health policy circles, but data repositories derived from human subjects may have to operate under managed access, to protect privacy, align with participant consent and ensure that the resource can be managed in a sustainable way. Data access committees need to be flexible, to cope with changing technology and opportunities and threats from the wider data sharing environment.

### Trade-off between timeliness and accessibility

The spread of SARS-CoV-2 has taught us that prevention relies on community engagement and has exposed the fragility in our research relationships. In an ideal scenario, health workers, such as doctors, virologists, data custodians at data-repositories, biobanks and in pharma, as well as students at the university should be able to quickly collect, share, and analyze health data in a global, cross-border manner without complicated, lengthy access-procedures. The ability to share data securely should not be restricted to selected specialists in the field but should be possible for all healthcare workers, as such the access and sharing procedures need to be easy. Data collected during a pandemic should not only be accessible, but also analyzable; meaning all stakeholders and the public should have the ability to analyze the data, not only the data custodians or project leads of research consortia.

Having such an affordable and easy-to-use platform in place, even developing countries are enabled to work together to reduce a pathogens’ spread and minimize its impact, despite severe resource constraints. However, our health systems are -more often than not-conservative and innovation averse.

What is needed is true leadership in the research community that is able to adopt new solutions, who are ready to tackle the uncertainty a situation like COVID-19 creates. There is no value in competing for data during a global crisis, instead humanity needs to collaborate more than ever before to defeat a common enemy that has taken the world hostage. Collective solutions that provide a ‘one-stop shop’ for the centralization of information on pathogen <> host interactions can help ensure that appropriate conditions for collaborative research and sharing of preliminary research findings and data are in place to reap their full benefits.

The provided platform offers such a one-stop shop solution, without being an exclusive storing solution. Data shared on the platform can also be collected and shared elsewhere. Beyond short-term policy responses to COVID-19, the Shivom platform supports research and innovation that could help tackle future epidemics early, on a national or international scale.

## Methods

The Shivom data analytics platform was built on a principle of open science, transparency, and collaboration and has been designed to give researchers a one-stop ecosystem to easily, securely and rapidly share and analyze their omics data, therefore simplifying data acquisition and data analysis processes. No coding proficiency is needed for using the platform, so everyone can use it and get results, minimizing work times. Our organization started out with the mission to accelerate data access, easy analysis, and exchange across international and organizational boundaries to promote equal access to healthcare for all.

The platform facilitates sharing and analysis of genetic, biomedical, and health-related data in a safe and trusted environment and contains two connected marketplaces, one for omics data and one for bioinformatics/AI pipelines and tools (see Fig. 1).

**Figure 1:**
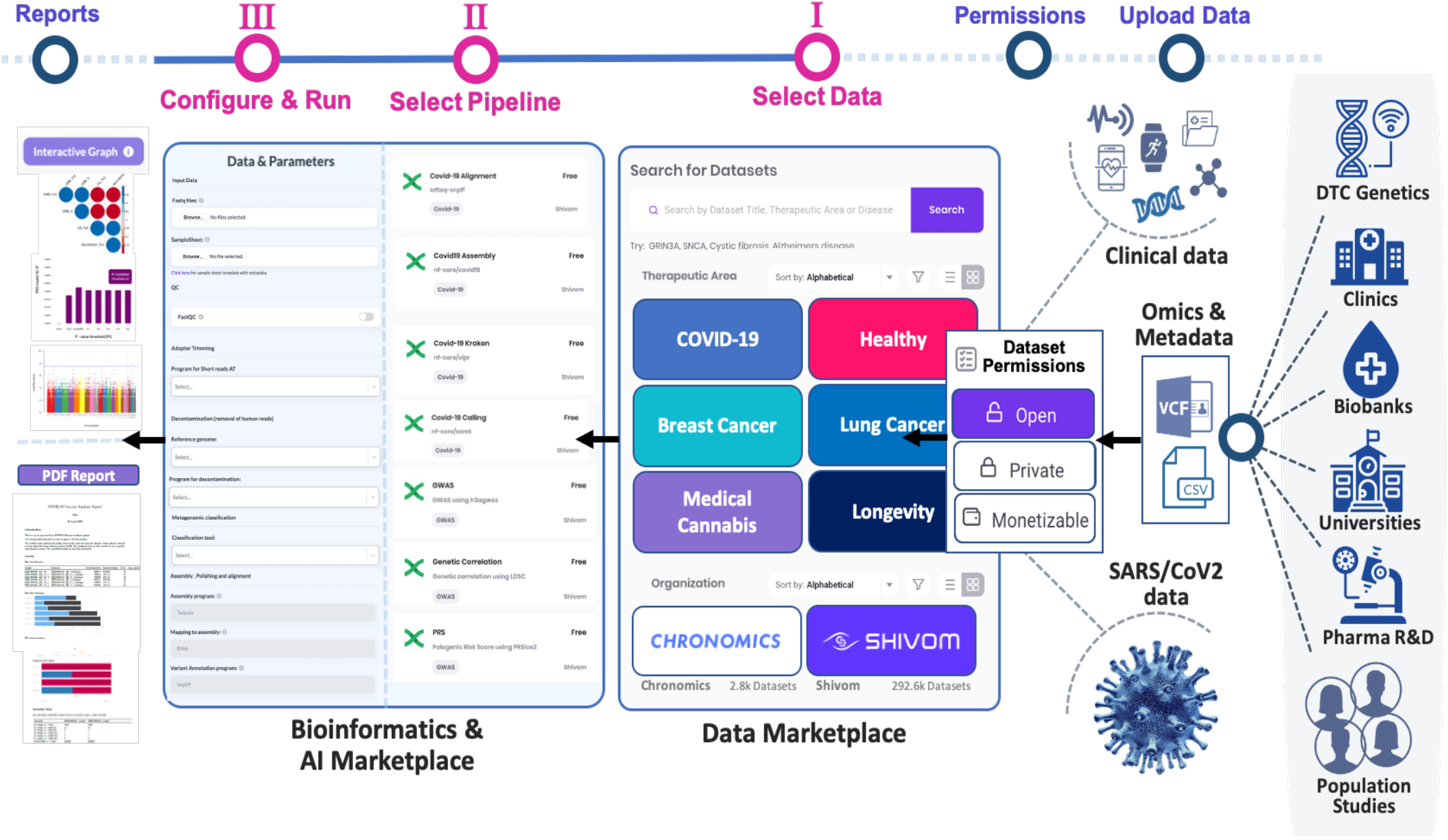
Data Flow in the Shivom platform. The user does not need in-depth bioinformatics expertise and is guided through all necessary steps from data upload to data analytics. Basically, data from a variety of sources is uploaded by the data custodian and data permission rules are set. If the data is set to open or monetizable, it will be searchable in the data marketplace. Once a dataset of interest is identified and filtered, the user can select a bioinformatic workflow (pipeline), configure user-specific parameters if needed, and then start the analysis. The user can then investigate and share the resulting report and, depending on the workflow, run further downstream analysis.

It was shown recently that clinical data collection with rapid sequencing of patients is a valuable tool to investigate suspected health-care associated COVID-19 cases^15^. Similarly, the Shivom platform can enable quick insight into local outbreaks to identify opportunities to target infection-control interventions to further reduce healthcare associated infections. If widely adopted by researchers and doctors around the globe, this wealth of genomic and healthcare information is a powerful tool to study, predict, diagnose and guide the treatment of COVID-19. The information from combined epidemiological and genomic investigations can be fed back to the clinical, infection control, and hospital management teams to trigger further investigations and measures to control the pandemic.

### Access Management

Signing up on the platform is easy and doesn’t require complicated access procedures. The idea behind this mechanism is to provide access to clinical and health data and analytics tools for everyone. Calls for data sharing have mostly been restricted to publicly funded research, but we and others argue that the distinction between publicly funded and industry-funded research is an artificial and irrelevant one^16^. At the center of precision medicine should be the patient which should override commercial interests and regulatory hurdles. Data sharing would lead to tremendous benefits for patients, progress in science, and rational use of healthcare resources based on evidence. As such, the Shivom platform stands out for its ease of use from the very first steps; the only requirements are a valid email, preferably institutional, in case the user wants to use advanced features that come with institutional plans. The user needs to set a password, and if required, can utilize an additional 2 step verification procedure to add an extra layer of security.

The personal data stored by the platform is protected under GDPR regulations whether it is hosted in the European Union or another country on other continents. After the sign-up, users are directed to a settings and profile page that allows to add additional information about the user and organization, research interests and dataset needs, or to switch between light and dark mode to make the user experience more personalized. The personal profile can be changed at any time.

Depending on the account type (e.g. academic group or company), the user can invite team members to the workspace to work on projects together and to share data and bioinformatics pipelines. Team members can be easily assigned user roles, i.e. any new user can be upgraded to administrator for the account.

Team members can be removed any time from the account by the administrator (project lead or manager). With the ‘Manage Plan’ tab, the platform can be adapted and scaled to meet the requirements of the users, from single/private users (e.g. students or freelance bioinformaticians), academia and startups, to small and medium enterprises with vast amounts of data and unique requirements as well as pharmaceutical organizations looking for custom solutions.

### Data Management

The Shivom platform was designed to make the process of uploading and sharing COVID-19 and other genetic data as easy and simple as possible. The reason is that access to the data sharing and analysis features should not be restricted to a few trained experts (data analysts or bioinformaticians) but should be easy for all healthcare professionals. Importing records (genetic or metadata files) into the user’s workspace is done via the ‘*My Datasets*’ page. In case of sequencing data, the user will primarily choose to upload Variant Call Format (VCF) files. VCF is a tab delimited text file format, often stored in a compressed manner, that contains meta-information lines, a header, and then data lines each containing information about a position in the genome^17^. The format also has the ability to contain genotype information on samples for each position in the genome (virus or host).

VCF is the preferred format on the platform because it is unambiguous, scalable and flexible, allowing extra information to be added to the info field. Many millions of variants can be stored in a single VCF file, and most bioinformatics pipelines on the platform use VCF files as input. Most NGS pipelines use the FASTQ file format, the most widely used format, generally delivered from a sequencer. The FASTQ format is a text-based format for storing both a nucleotide sequence and its corresponding quality scores. FASTQ.gz is the preferred file format for the standard COVID-19 pipelines preinstalled on the Shivom platform.

Despite VCF being very popular especially in the statistical genetic area, there are also other file formats that can be used on the upload or directly at the point of configuring pipeline parameters. The purpose of these other genetic file formats is to reduce file size – from many VCFs to just three distinct files: a) .bed (Plink binary biallelic genotype table), that contains the genotype call at biallelic variants, b) .bim (Plink extended MAP file) which also includes the names of the alleles: (chromosome, SNP, cM, base-position, allele 1, allele 2); c) .fam (Plink sample information file), is the last of the files and contains all the details with regards to the individuals including, whether there are parents in the datasets, and the sex (male/female). This file will also contain information on the phenotype/traits of the individuals in the cohort.

Ideally, the sequencing file is accompanied by phenotypic metadata (e.g. is it patient or control data, age, height, comorbidities, etc..), describing the uploaded datasets (see Metadata). The metadata file is uploaded as a CSV file, a comma-separated values file, which allows data to be saved in a tabular format. CSV files can be used with most any spreadsheet program, such as Microsoft Excel or Google Spreadsheets. For ease-of-use, a sample template file is provided on the platform.

### Data Marketplace Permission Settings

In the next step, the user has the option to save specific data sharing permission settings related to the uploaded dataset to allow for fine-grained data sharing with others. These unique permission settings transform the data analytics platform into a massive data marketplace. Providing health information for a data marketplace and sharing it is a key in leveraging the full potential of data in precision medicine and the fight against pandemics.

The marketplace is a central point of entry for the users in order to find needed data faster and more structured. In addition, interoperability can be created through the marketplace, which supports cross-sectoral applications. Unrestricted access to relevant pandemic data in particular will become increasingly important in the future.

The user has three options:

**Open –** All data designated open will be available for the public to analyze. However, open does not mean that the data is downloadable, this permission setting only allows only for algorithms/pipelines to use the data in the data analytics module. All open data is searchable on the platform. Being a data-hub for COVID-19 data, we highly encourage all researchers and data custodians to make their data open/public and accessible for the scientific community, thereby increasing the amount of open-source COVID-19 data currently available and usable.

**Private –** Private data will be stored in a secure, private environment, and is only available and analyzable by the data owner and team members from the same organization or project and will not show up in data search results. This setting is usually used when data is not supposed to be shared (or only within a consortium), or if the data needs to be analyzed or published before sharing commences.

**Monetizable –** By choosing this option, the data owner/custodian authorizes the marketplace to license their information on their behalf following defined terms and conditions. In this context, data consumers can play a dual role by providing data back to the marketplace. Data marketplaces make it possible to share and monetize different types of information to create incremental value. By combining information and analytical models and structures to generate incentives for data suppliers, more participants will deliver data to the platform and the precision medicine ecosystem. To monetize pandemic data, making datasets available for researchers at a cost, can make sense if a laboratory is not otherwise able to finance the data collection, e.g. when located in an underfunded developing nation.

At any time, the permission settings of data files can be updated. The settings have an audit trail and the platform owner or server host are not able to change the settings, only the data owner/custodian is able to modify the settings.

In the next step during data import is to add additional information to the dataset, such a dataset name, a Digital Object Identifier (DOI), health status, Author or Company name, a description of the dataset, or the name of the genotyping or sequencing system used (a list of sequencing platforms is available in a drop down menu). Other features will be added in later versions.

After requesting the user’s digital signature to confirm the operations carried out so far, the data will be uploaded, and the user will be redirected to a page containing a summary of his/her data sets. During the upload, the data is checked for quality metrics and for duplicates (compared to data already existing on the data marketplace) by internal algorithms that prevent monetization of datasets that were shared by others before. Genetic data is largely transformed into a common format as part of the data curation process. The platform also starts automated proprietary anti-fraud mechanisms to identify data that was tampered with.

All data in a user’s workspace are easily searchable and can be sorted by therapeutic area (based on collection of biomedical ontologies) and other parameters. Generally, the workspace allows for building larger datasets in time, and at any point to share data whenever it is needed, or to revoke access. There are several potential use cases for using the platform as storage space for data.

First of all, having a sharing infrastructure will enable healthcare professionals to quickly accumulate and share anonymized data with the public (e.g. sequencing data from COVID-19 patients), which is particularly important during an early disease outbreak.

Second, the platform is an ideal storage infrastructure for data custodians (e.g. biobanks, direct to consumer genetic companies, or patient support groups) that inherently have large curated patient and volunteer datasets. The data can then be monetized, but more importantly, the datasets can be made available to the wider research and scientific community to make medical breakthroughs. Since the basis of the platform is precision medicine, at this point in time, data custodians are encouraged to store their data as VCF files; however, a plethora of other file formats are planned to be added to the platform in time. VCF are particularly useful as they are one of the final file formats of most genomic technologies across both microarray chips and sequencing (genome VCF/ gVCF). They are also the most common file format when it comes to secondary genetic analysis, but equally their smaller file size reduces the cost on storage.

Third, using the platform as a repository for research data during a peer-review publication process. Researchers are increasingly encouraged, or even mandated, to make their research data available, accessible, discoverable and usable. For example, the explicit permission settings in the platform would allow researchers to grant access to the journal’s reviewers to their data and analyses, prior to publication. This time-restricted data sharing can be valuable when data should not yet be available to the wider public.

Fourth, the platform is an ideal data sharing infrastructure for research consortia. Groups that are located in different countries can easily share data and result and build larger, statistically more relevant datasets. Such a procedure is particularly valuable for pre-competitive consortia, to significantly lower the costs to accumulate valuable datasets, while the in-house research can be kept confidential.

### Patient privacy & data security

The Shivom platform was developed with a comprehensive security strategy to protect user data privacy and security.

### Patient Privacy

The central pillar of the Shivom data-hub is patient data privacy and confidentiality. The duty of confidentiality is a medical cornerstone since the Hippocratic Oath, dating back to the 5^th^ century BC. On the other hand, the right to privacy is a relatively recent juridical concept^18^. The emergence of genetic databases, direct-to-consumer genetics, and electronic medical record keeping has led to increased interest in analyzing historical patient and control data for research purposes and drug development. Such research use of patient data, however, raises concerns about confidentiality and institutional liability^19^. Obviously, doctors and other healthcare workers have to make sure that no private and protected information of a COVID-19 patient, such as name, address, phone number, email address, or biometric identifiers are exposed to 3^rd^ parties. As such, institutional review boards and data marketplaces must balance patient data security with a researcher’s ability to explore potentially important clinical, environmental and socioeconomic relationships.

In this context, the platform was designed from the beginning to be GDPR compliant and to provide full anonymization of patient records (privacy by design). The advantage of this approach is that personal or confidential data that has been rendered anonymous in such a way that the individual is not or no longer identifiable is no longer considered personal data. As such, by making it impossible to connect personal data to an identifiable person, data controllers (i.e. the data custodian or patient who can upload and manage data) and processors (researchers) are permitted to use, process and publish personal information in just about any way that they choose. Of course, for data to be truly anonymized, the anonymization must be irreversible. Anonymization will provide opportunities for data controllers to use personal data in more innovative ways for the greater good. Full anonymization is achieved with several steps. First of all, no private, identifiable data of patients is stored together with clinical data. Second, and most importantly, the platform provides only summary statistics/analytics on the data. That means sharing of data only allows bioinformatics pipelines or machine learning algorithms to touch the data; no raw data (e.g. DNA sequencing reads) can be downloaded or viewed by the data user/processor.

All data must be stripped of any information that could lead to identification, either directly or indirectly—for instance, by combining data with other available information, such as genealogy databases^20^. However, at this point in time, the consent mechanisms as well as the process of removing private or confidential information from raw data remains with the data custodian, for example the hospital that uploads the COVID-19-related sequences. In this context, it is important to understand that only the data owner/custodian has all the access rights and can manage the data such as granting permissions to use the data. The platform provider has no power to manage the data and only has the possibility to identify duplicate datasets that will automatically be excluded from the data marketplace.

Other security features on the platform are that no statistical analyses are possible on single individuals, and only vetted algorithms and bioinformatics pipelines are available on the analytics marketplace to avoid that algorithms are deployed to specifically read out raw data from individuals or to cross-link data from individuals to other, de-identifying databases.

### Data Security & Encryption

By harnessing the power of cloud computing the platform enables data analysts and healthcare professionals, with limited or no computational and data analysis training, to analyze and make sense of genomic data and COVID-19 in the fastest and most inexpensive way possible. The genetic data stored with the platform is handled and managed with state-of-the-art technology and processes, using a combination of security layers that keep data and management information (cryptographic keys, permission settings etc.) safe. Currently, all the COVID-19 related data is uploaded to Amazon Web Services (AWS) S3 buckets that provide data storage with high durability and availability. Like other services, S3 denies access from most sources by default.

The Shivom platform requires specific permission to be allowed to perform transactions or computation actions on the bucket. The S3 storage service allows to devote a bucket to each individual application, group or even an individual user doing work. Access to the buckets is regulated via the platform on different, non-AWS server(s), and all information related to the access is encrypted using state-of-the-art Key Management Services (KMS). In addition, no user/custodian data is stored alongside genetic data and is completely separated and encrypted using different cloud storage solutions.

### Immutable Audit Trail & Smart Contracts

One powerful feature of our solutions is the ability to maintain an immutable data-access audit trail via blockchain. This immutability property is crucial in scenarios where auditability is desired, such as in maintaining access logs for sensitive healthcare, genetic and biometric data. In precision medicine, access audit trails for logs provide guarantees that as multiple parties (e.g., researchers, laboratories, hospitals, insurance companies, patients and data custodians) access this data create an immutable record for all queries^21–23^. Since blockchains are distributed, this means that no individual party can change or manipulate logs^24,25^. Such a feature can help journal reviewers and research collaborators to protect themselves from inflation bias, also known as “p-hacking” or “selective reporting”. Such bias can be the cause of misreporting or misinterpretation of true effect sizes in published studies and occurs when researchers try out several statistical analyses and/or switch between various data sets and eligibility specifications, and then selectively report only those that produce significant results.

All permission settings stored in the system are coded into a blockchain, guaranteeing that nobody can temper with the permission setting provided by the data owner. Each data transfer on the platform represents a cryptographic transaction hash. Hashing means taking an input string from the platform of any length and giving out an output of a fixed length. In the context of blockchain technology, the transactions are taken as input and run through a hashing algorithm (e.g. a hash function belonging to the Keccak family, the same family which the SHA-3 hash functions belong to) which gives an output of a fixed length that is stored immutable on the chain.

A transaction is always cryptographically signed by the system. This makes it straightforward to guard access to specific modifications of the database. Transactions such as monetization of datasets are encoded as smart contracts on the blockchain. A “smart contract” is simply a piece of code that is running on a distributed ledger, currently Ethereum. It’s called a “contract” because code that runs on the blockchain can control digital assets and instructions. Practically speaking, each contract is an immutable computer program that runs deterministically in the context of a Virtual Machine as part of the Ethereum network protocol—i.e., on the decentralized Ethereum world computer. Once deployed, the code of a smart contract cannot change. Unlike with traditional software, the only way to modify a smart contract is to deploy a new instance, making it immutable.

All the contracts used by the Shivom platform only run if they are called by a transaction, e.g. when a file is used in a data analytics pipeline. In this case, a smart contract is triggered that can, if the file permissions are set to monetizable, transfer a fee to the electronic wallet of the data owner. The outcome of the execution of the smart contract is the same for everyone who runs it, given the context of the transaction that initiated its execution and the state of the blockchain at the moment of execution.

If needed, smart contracts can be put in place to accommodate specific terms needed in more complex research or drug development agreements, e.g. if datasets belong to a consortium or a biobank, or if there are data lock constraints (e.g. an embargo on data release) and usual access procedures are complicated. For example, most biobanks contracts for accessing data are long, arduous, detailed documents with complicated language. They require lawyers and consultants to frame them, to decipher them, and if need be, to defend them. Those processes can be made much simpler by applying self-executing, self-enforcing contracts, providing a tremendous opportunity for use in any research field that relies on data to drive transactions. These contracts already possess multiple advantages over traditional arrangements, including accuracy and transparency.

The terms and conditions of these contracts are fully visible and accessible to all relevant parties. There is no way to dispute them once the contract is established, which facilitates total transparency of the transaction to all concerned parties. Other advantages are speed (execution in almost real-time), security (the highest level of data encryption is used), efficiency, and foremost trust, as the transparent, autonomous, and secure nature of the agreement removes any possibility of manipulation, bias, or error.

### Metadata

Advances in data capture, diagnostics and sensor technologies are expanding the scope of personalized medicine beyond genotypes, providing new opportunities for developing richer and more dynamic multi-scale models of individual health. Collecting, and sharing phenotypic and socioeconomic data from patients and healthy controls, in combination with their genotype and other omics profiles, present new opportunities for mapping and exploring the critical yet poorly characterized “phenome” and “envirome” dimensions of personalized medicine. Therefore, in the context of COVID-19, if available, metadata should be shared next to sequencing information of the patients. Such metadata can contain information with regards to the comorbidities, ethnic background, treatment and support that they receive rather than biological or clinical data. Currently, metadata is uploaded as a CSV file to the platform, which allows almost all phenotypic data to be saved in a tabular format. Other inputs may be available in the future, e.g. to parse information directly from a medical record or from wearables. Using a simple tabular format also allows it to be source agnostic, e.g. it is not difficult to combine genetic information with patient questionnaires.

Indeed, patients support groups, direct-to-consumer genetic companies or patient registries are less likely to have biological data types in their repositories and often have only metadata that comes from questionnaires that volunteers have filled. An example of this is The Dementia Platform UK (DPUK)^26^, a patient registry that comprises thousands of individuals from across dozens independent cohorts - which allows a variety of analysis to be conducted including machine learning and mendelian randomization techniques. The platform is multi-purpose built, which allows for all types of data that is alpha-numerical encoded to be stored. Other platforms are engineered specifically for a particular type of cohort, e.g. tailored towards analyzing biobank data. The Shivom platform is completely source agnostic and is engineered to allow large cohort datasets to be uploaded regardless of biobanks or cohorts. Additionally, the platform will allow for biostatistical (non-genetic), epidemiological and machine learning analysis types to be undertaken.

The metadata is structured in a simple and intuitive way in order to allow the user to be able to quickly query the dataset. Depending on whether the data being uploaded has accompanying genetics information, the first column of the file will either contain the name of the VCF file or the first metadata attribute. CSV files are supported by nearly all data upload interfaces and they are easier to manage if data owners are planning to move data between platforms, export and import it from one interface to another. A template that illustrates how metadata can be organized, including a standard metadata set (e.g. height, smoking status, gender, etc..) is provided on the platform.

For COVID-19 datasets (provided as VCF) we encourage data providers to share as many as possible phenotypic, clinical and socioeconomic data, keeping in mind that all data should be anonymized. Working with several COVID-19 consortia, we established a guideline of what an ideal dataset should include, keeping in mind that many clinical data are usually not available 9see supplement for metadata structure):

- Ethnicity, Age, Gender, Height, Weight
- Smoking information
- Comorbidities (i.e. Diabetes, cardiovascular diseases, renal insufficiency, COPD, Malignant tumor, autoimmune diseases, immunodeficiency, asthma, cystic fibrosis, Dementia, Liver disease, Hemiplegia, Leukemia, Lymphoma, AIDS…)
- Charlson comorbidity index^27^
- Pneumonia status: non-present, present: unilateral, present: bilateral (dual)
- Sequential Organ Failure Assessment (SOFA) score^28^
- Clinical Pulmonary Infection Score (CPIS)^29^
- Quick sepsis related organ failure assessment (qSOFA)^30^
- Clinical Outcome: oxygen, continuous positive airway pressure (CPAP), intubation data, days with symptoms, antiviral at diagnosis, mortality, and clinical laboratory data.

### Data Search

The platform supports fine-grained search queries across global omics datasets. The idea is to find and analyze genetic and other omics dataset from various global sources in the easiest, fastest, and most efficient way. On the ‘*Discover*’ page, with just a few simple steps, the user will be able to quickly discover and select the data according to the disease of interest and filter the dataset based on the appropriate metadata for their study or research question.

One strength of the platform is that the user can find and combine open-source, in-house, and proprietary datasets in a matter of seconds. The user has the opportunity to perform a free search with advanced search operators (e.g. searching for gene- or disease-name), or can use separate clinical search bars, and therapeutic area, disease or dataset title. The Discover window will then be populated with the search results. Currently, the search query form supports DOID ontology, a comprehensive hierarchical controlled vocabulary for human disease representation. but other standards (EFO, ICD10) are planned to be added in the future as well.

### Stratifying Patients

Once the data has been selected, it will be possible to further filter it based on the information contained in the associated metadata files, generally gender, diet, country, etc., and also the type of data permission associated. For example, it is possible to select only free data that is not associated with any costs. This fine-grained filtering feature therefore allows to stratify patients based on the information the user is interested in, so that it will be possible to select the most significant data for the analyzes to be conducted. For example, once the database is populated, it should be possible to select COVID-19 patients by ethnicity or smoking status to quickly find confounding factors that may affect the outcome and severity of the disease.

Once the user has completed the selection of the data that has to go into the data analysis, a simple click on the ‘Choose Pipeline’ box will bring the user to the data analytics marketplace for downstream analytics.

### Data Analytics Marketplace

After selecting datasets for analysis, the user is directed to the data analytics marketplace. The marketplace is designed to provide researchers with or without bioinformatics expertise to perform a wide variety of data analyses and to share in-house pipelines as well as machine learning tools with the scientific community. Since the launch of the platform, a variety of standard bioinformatics pipelines/workflows, primarily for secondary analyses, were already added to the marketplace that are free to use, including pipelines to analyze COVID-19 patients. The Shivom analytical platform hosts a variety of genomic-based pipelines; from raw assembly and variant calling, to statistical association-based analysis for population-based studies. In time, a vast variety of new data analytics tools and pipelines will be added to the marketplace, including AI analytics features.

Currently, all provided standard pipelines are built on Nextflow technology^31^. Nextflow is a domain-specific language that enables rapid pipeline development through the adaptation of existing pipelines written in any scripting language^32^. Nextflow technology was chosen for the platform because it is increasingly becoming the standard for pipeline development. Its multi-scale containerization makes it possible to bundle entire pipelines, subcomponents and individual tools into their own containers, which is essential for numerical stability. Nextflow can use any data structure and outputs are not limited to files but can also include in-memory values and data objects^32^. Another key specification of Nextflow is its integration with software repositories (including GitHub and BitBucket) and its native support for various cloud systems which provides rapid computation and effective scaling.

Nextflow is designed specifically for bioinformaticians familiar with programming. However, it was our aim to provide a platform that can be used by inexperienced users and without common line input. As such, the Shivom data analytics marketplace was designed to provide an easy to use graphical user interface (GUI) where all pipelines can be easily selected and configured, so that no coding experience is required by the user. In addition to the data marketplace, this feature sets the platform apart from other cloud computing tools that use Nextflow or other workflow management tools such as Toil, Snakemake or Bpipe. Users can easily automate and standardize analyses (e.g. GWAS, Meta-GWAS, Genetic Correlation, Two Sample Mendelian Randomization, Polygenic Risk Score, PheWAS, Colocalization etc..) by selecting all parameters for sets of different analysis activities. This makes it possible to run analyses fully automated, meaning that even new users can start using sophisticated analyses by running a workflow in which all analysis steps are incorporated, i.e. assembled by more experienced bioinformaticians or machine learning experts.

Findings that cannot be reproduced pose risks to pharmaceutical R&D, causing both delays and significantly higher costs of drug development, and also jeopardize the public trust in science (36). By using standard pipelines and allow for 3rd parties to share their workflows on the platform, and by creating reproducible and shareable pipelines using the public Nextflow workflow framework, not only do bioinformaticians have to spend less time reinventing the wheel but also do we get closer to the goal of making science reproducible.

### GWAS pipeline to study virus-host interactions

The disadvantage of many research consortia, including some COVID-19 consortia, is that they do not open the data to all consortium members to analyze the collected data individually, but only provide data analysis by a central coordinator. Then the summary statistics of the analyzed cohorts are fed back to the consortium members. For example, the Host Genetics Initiative agreed on a standardized Genome-wide association studies (GWAS) pipeline^33^. The Shivom platform works differently, and all participants of a research consortium (and other 3rd parties if permitted) can analyze the datasets independently. To make GWAS analyses available to the whole research community, one of the most important preinstalled pipelines on the platform is a standard GWAS analysis workflow for data quality control (QC) and basic association testing. Genome-wide association studies are used to identify associations between single nucleotide polymorphisms (SNPs) and phenotypic traits^34^. GWAS aims to identify SNPs of which the allele frequencies vary systematically as a function of phenotypic trait values. Identification of trait-associated SNPs may reveal new insights into the biological mechanisms underlying these phenotypes. GWAS are of particular interest to researchers that study virus-host interactions.

The SARS-CoV-2 virus does not affect everyone in the same way. Some groups seem particularly vulnerable to severe COVID-19, notably those with existing health conditions or with genetic predispositions. Variation in susceptibility to infectious disease and its consequences depends on environmental factors, but also genetic differences, highlighting the need for COVID-19 host genetics to engage with questions related to the role of genetic susceptibility factors in creating potential inequalities in the ability to work or access public space, stigma, and inequalities in the quality and scope of data^35^.

### COVID-19 pipelines

The genome sequencing of the coronavirus can provide clues regarding how the virus has evolved and variants within the genome. The highlighted platform contains a set of bioinformatics pipelines that allow the interrogation of host and virus genomes. Genetic information might enable targeting therapeutic interventions to those more likely to develop severe illness or protecting them from adverse reactions. In addition, information from those less susceptible to infection with SARS-CoV-2 may be valuable in identifying potential therapies.

The current version of the platform comes with preinstalled COVID-19 specific pipelines covering assembly statistics, alignment statistics, virus variant calling, and metagenomics analysis, and other pipelines that can be used for analyzing Covid-19 patient data such as GWAS or MetaGWAS. The research community is encouraged to add more pipelines and machine learning tools to support the fight against the pandemic. All the standard preinstalled COVID-19 pipelines work with FASTQ sequencing files. The platform works with Illumina short reads as well as Oxford Nanopore long reads. Long and short reads differ according to the length of reads produced and the underlying technology used for sequencing. The short-read length (100-200 bp) limits its capability to resolve complex regions with repetitive or heterozygous sequences, so the longer read lengths (up to kbs) have fundamentally more information. However, these tend to suffer from higher error rates than the short reads. The short reads come in zipped format (.gz) and do not need to be extracted before use; since there are paired-end, each sample will have two files: R1 and R2 FASTQ that need to be included in an analysis. Long reads come as single FASTQ files. Each analysis requires a sample metadata sheet, which specifies the FASTQ.gz files for each of the sample and other associated metadata files. A template metadata file can be accessed on the platform, in which the user can also find the description of all columns and the example values.

### Metagenomic Analysis

One of the common analyses done on patient sequences is to identify the pathogens contained in the analyzed sample and to assign taxonomic labels, usually obtained through meta-genomic studies^36^. Metagenomics allows researchers to create a picture of a patient’s pathogens without the need to isolate and culture individual organisms. The prediction power on COVID-19 also depends on data related to underlying morbidities, including other flu-like illnesses. Though metagenomic testing for other viruses can therefore increase the specificity of the GWAS studies and other algorithms.

The Shivom platform comes with a metagenomics analysis pipeline that is based on the nf-core/vipr pipeline, a bioinformatics best-practice analysis pipeline for assembly and intrahost low-frequency variant calling for pathogen samples. The pipeline uses the Kraken program metagenomics classification^37–39^. Using exact alignment of k-mers, the pipeline achieves classification significantly quicker to the fastest BLAST program.

The process of running pipelines on the platform are largely the same and do not require in-depth bioinformatics expertise or command line entry. The user is guided through all steps. After the upload of the selected FASTQ files and associated metadata, the user can choose to add a quality control and then to modify the parameters of the pipeline, if required.

After starting the computation, the user can easily monitor the progress of the pipeline and all other tasks in the ‘Projects’ tab (see Fig. 8).

**Figure 2:**
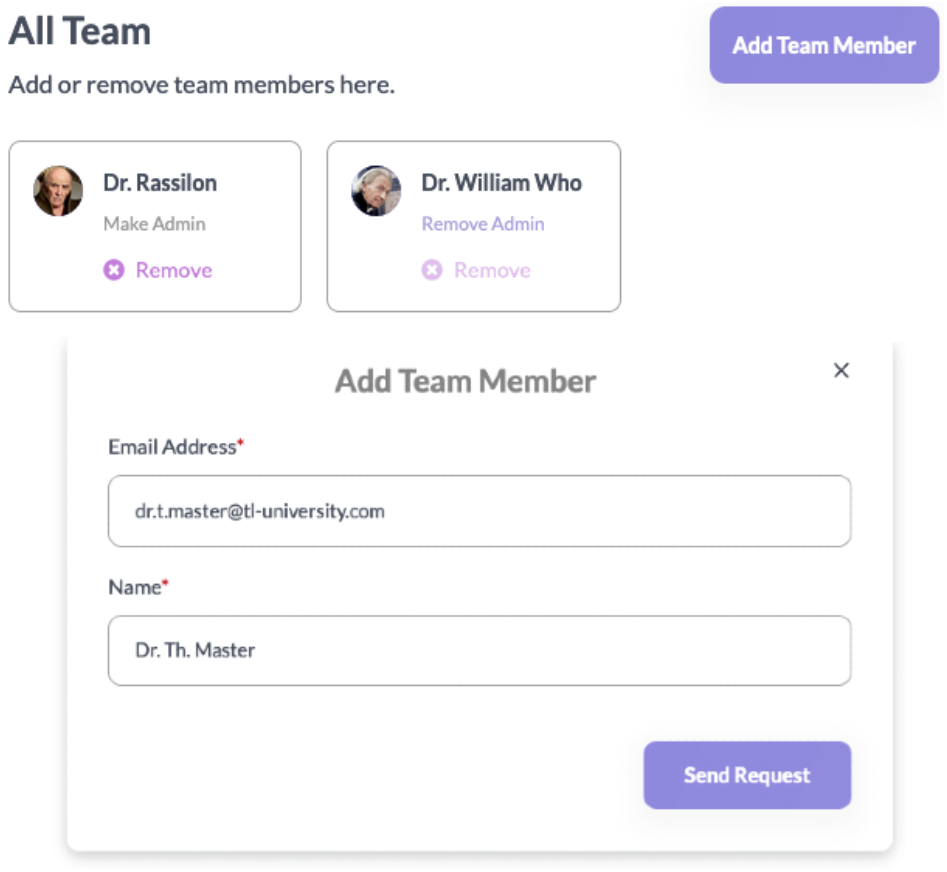
Team members from the same organization can be invited to join the workspace.

**Figure 3:**
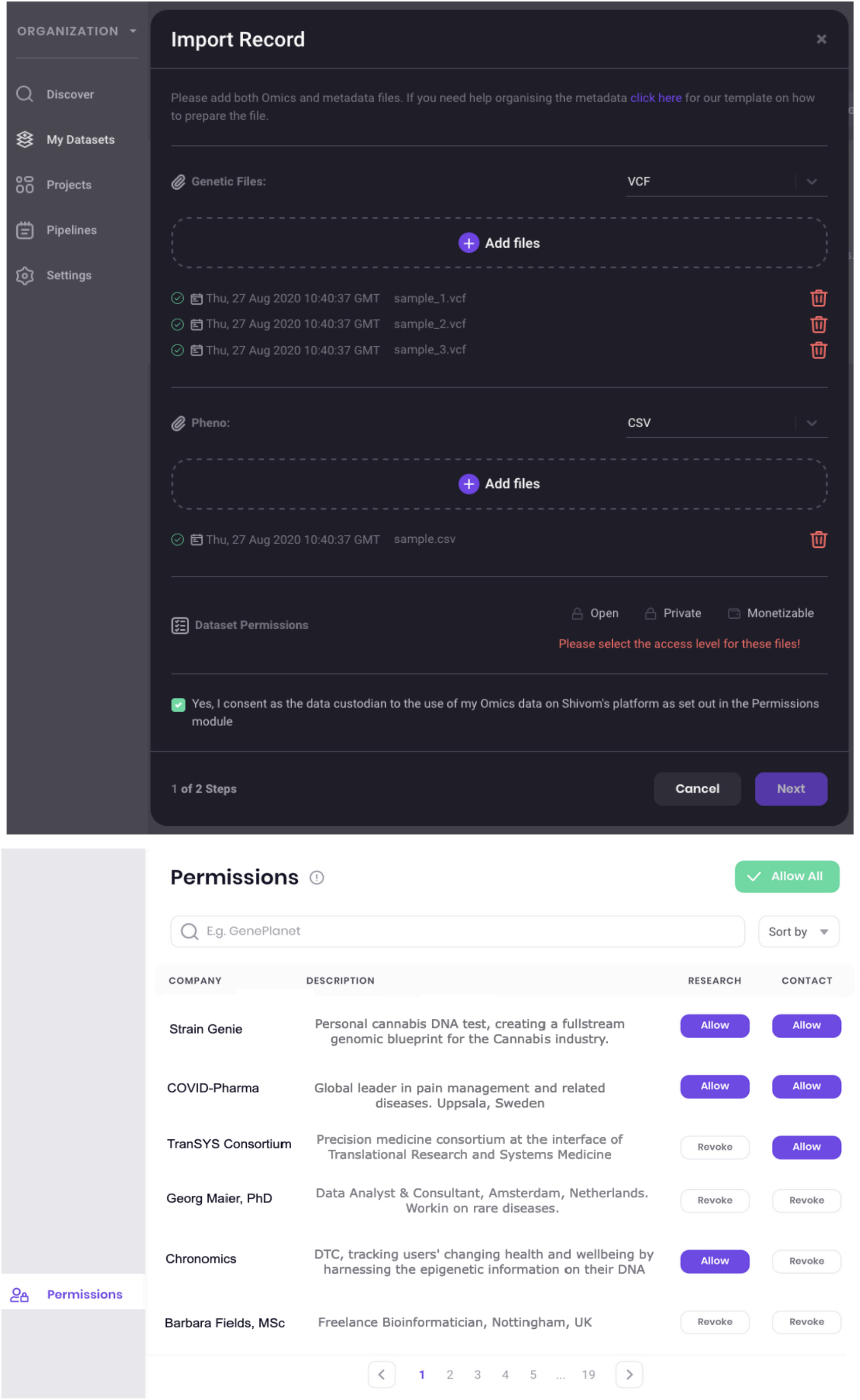
**Top:** Data Upload (dark mode). After choosing a file format from a drop-down menu, files can be either dragged and dropped to the workspace or uploaded via a new window. **Bottom**: Explicit data sharing permission are available to share data with selected collaborators, within a consortium, or with clients, co-workers, organizations or external analysts.

**Figure 4:**
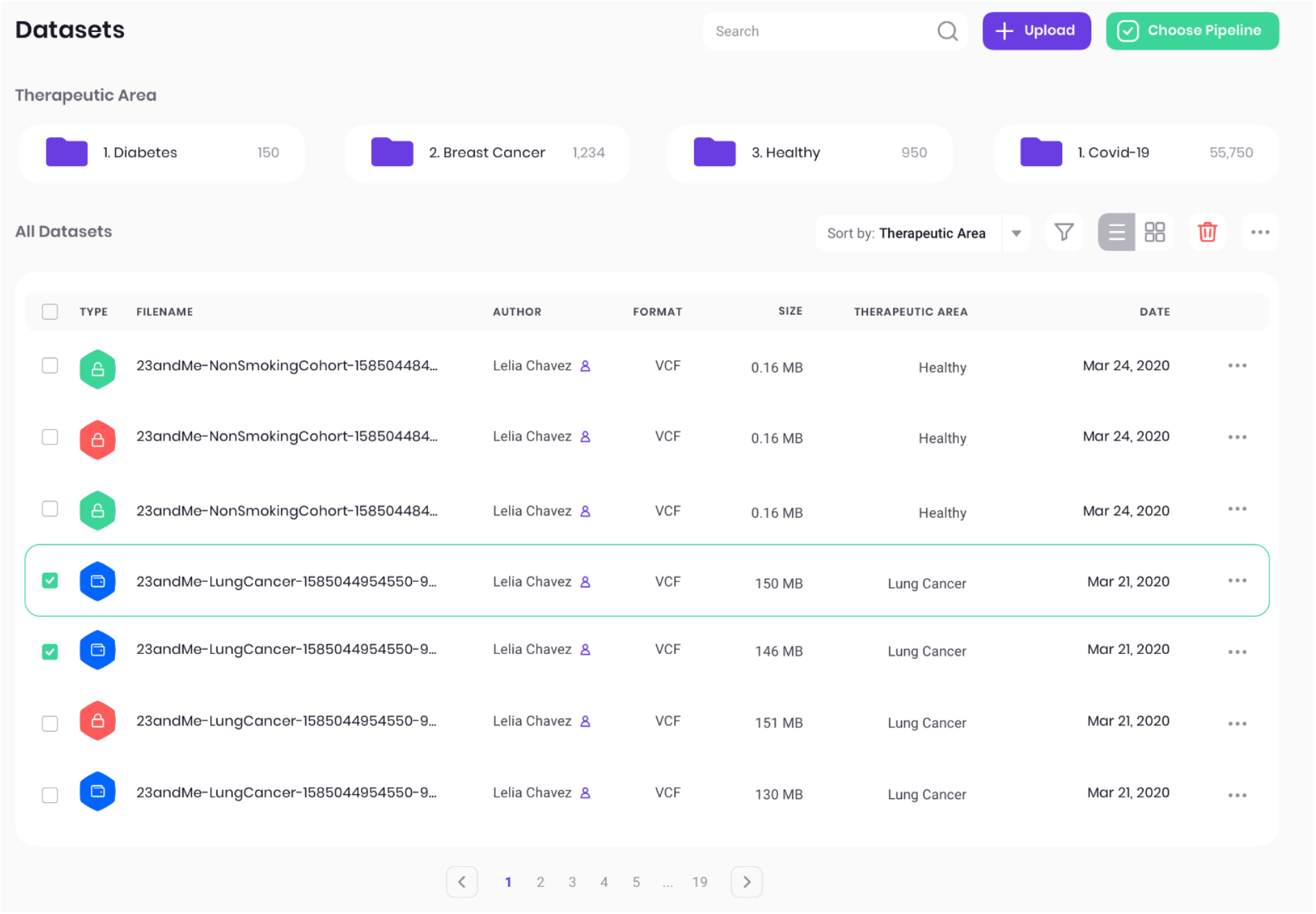
Dataset view that shows all data stored in the user’s workspace. The different permission settings are highlighted by designated icons and can be changed any time.

**Figure 5:**
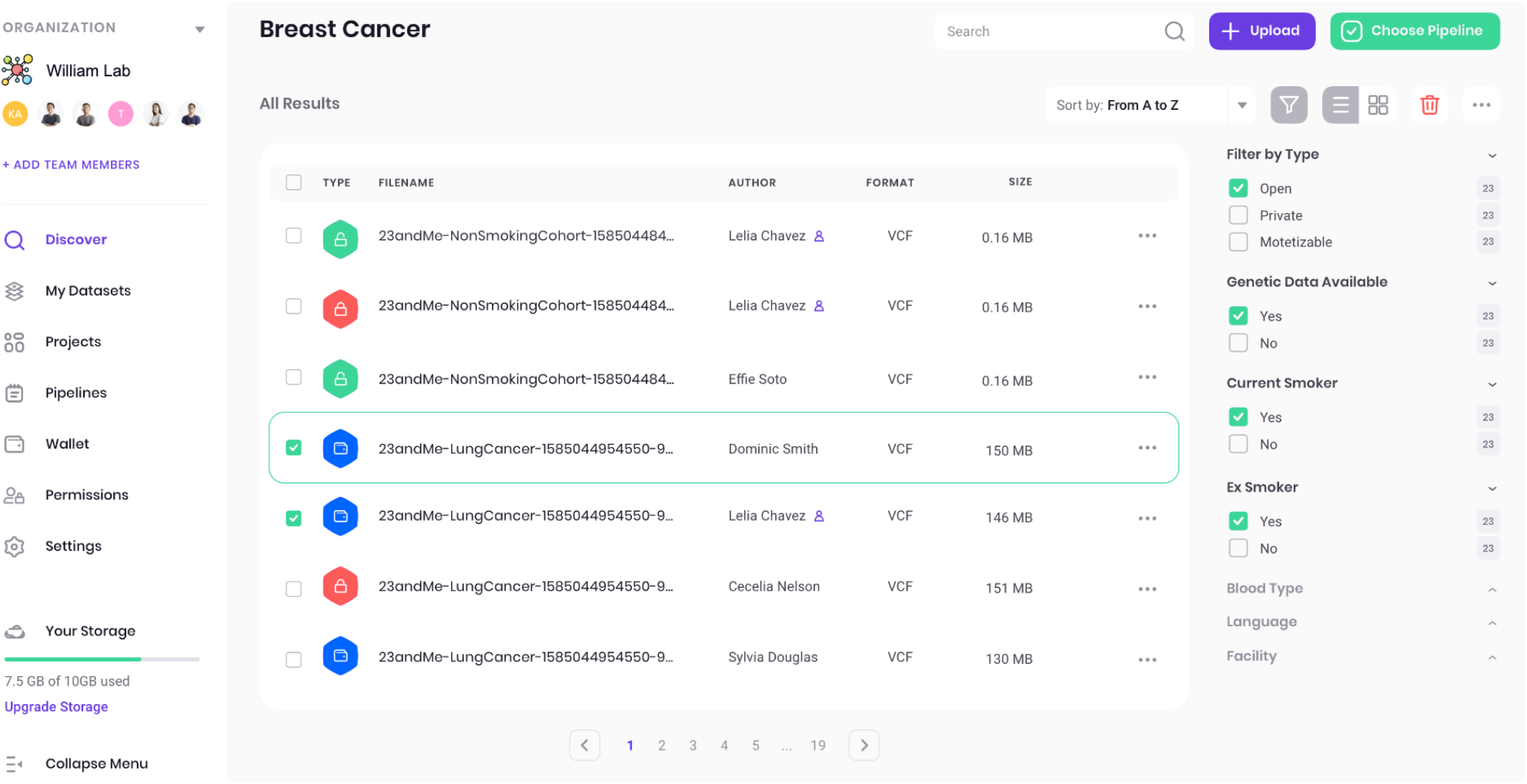
Discovery page with search results. On the right pane, the list of datasets can be further filtered in a fine-grained manner according to phenotypic or socioeconomic data contained in the associated metadata files, allowing for fine-tuning of the curated dataset.

**Figure 6:**
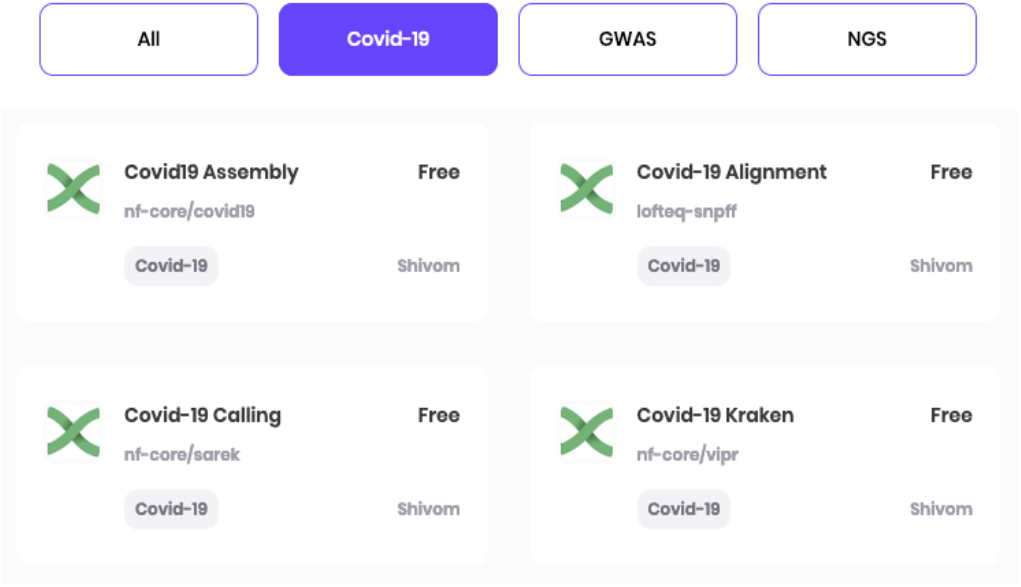
The data marketplace has a simple multiple-choice process: first, the user can choose which pipeline to use from a large variety of bioinformatics and AI pipelines, and can then define the parameters of the same.

**Figure 7:**
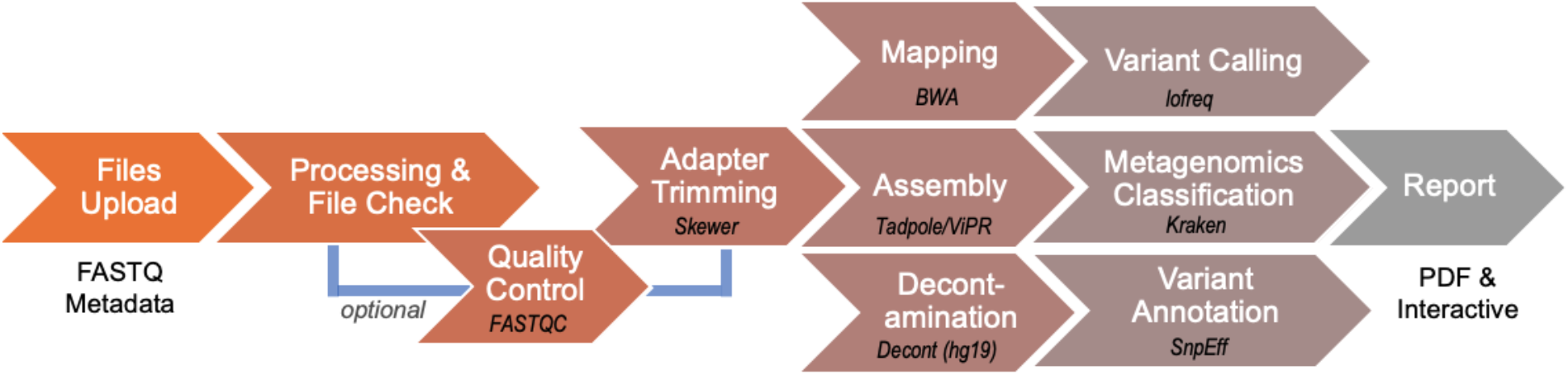
The Shivom COVID-19 Metagenomics Pipeline. The pipeline takes FASTQ (short- and long reads) together with metadata and will provide a comprehensive analytics report in PDF format and as interactive on-screen report.

**Figure 8:**
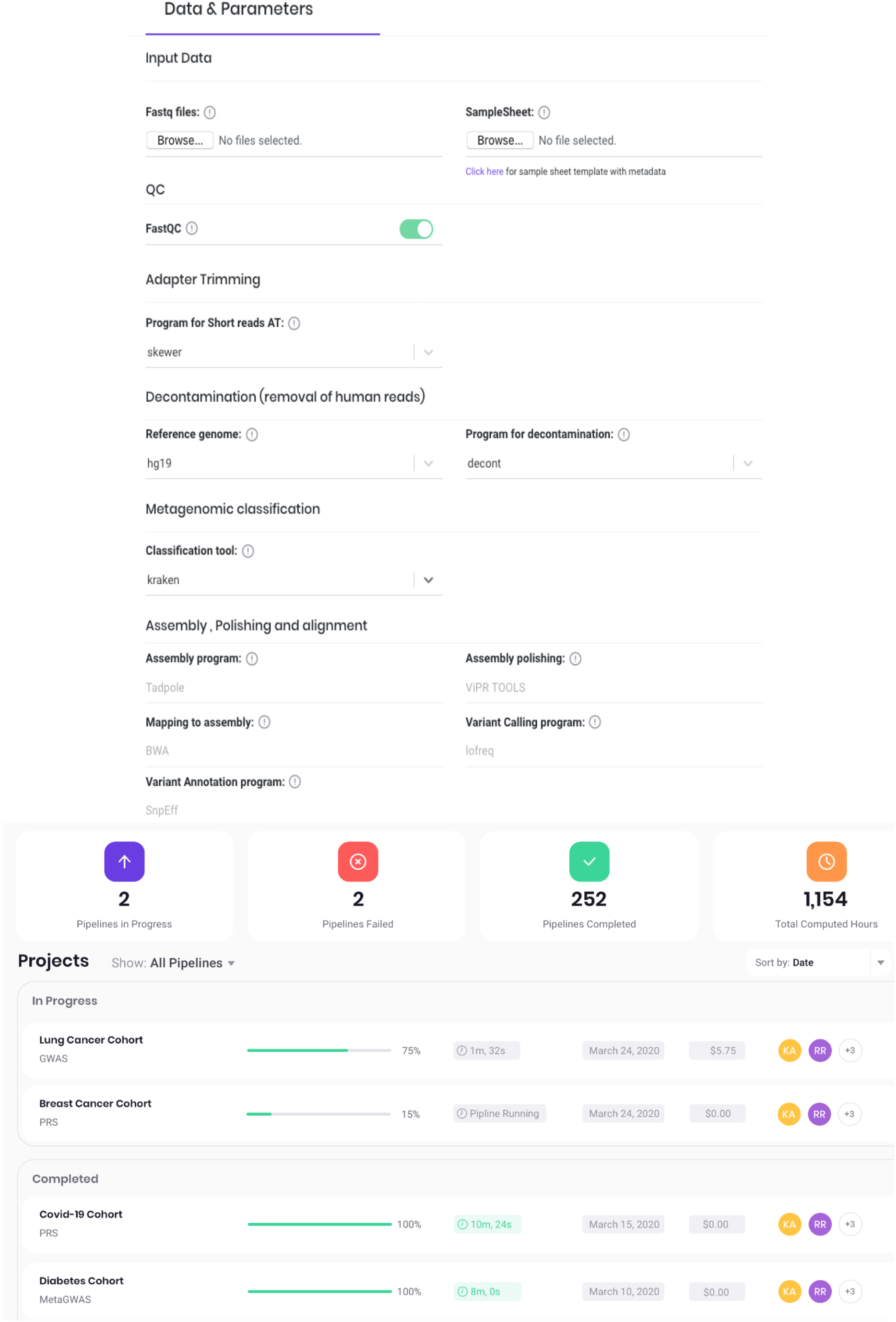
**Top:** Data & Parameters. After selecting the input files, the user can select and change the pipeline parameters, or can choose the default parameters recommended. The focus of the platform is on ease-of-use, no coding expertise is required. **Bottom:** The Projects view allows easy monitoring of all pipelines in progress and completed.

**Figure 9.**
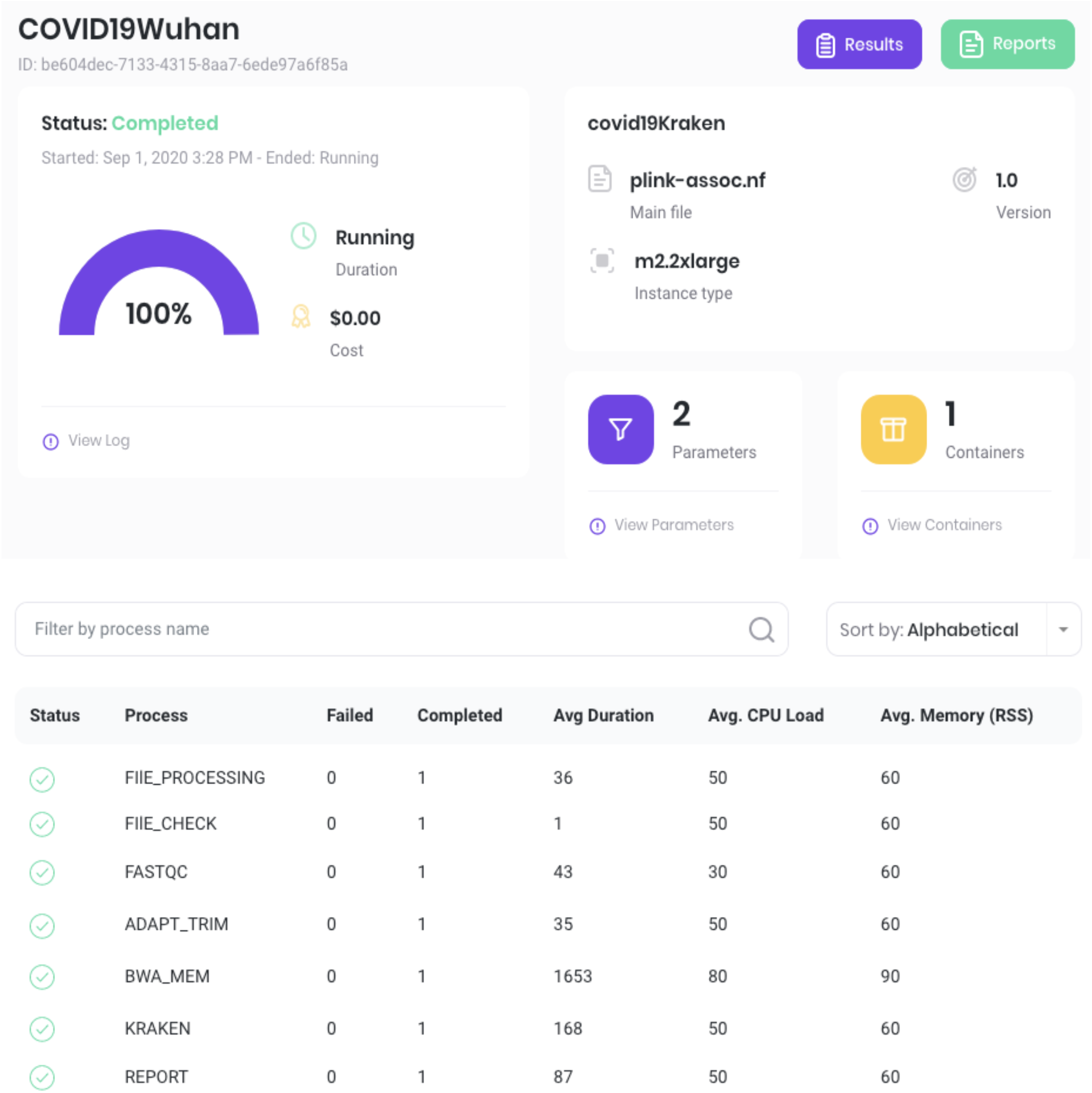
**Top:** The Pipeline view allows the user to monitor running and completed processes. **Bottom:** A list of all pipeline processes allows the user to monitor progress and helps in identifying potential errors.

**Figure 10:**
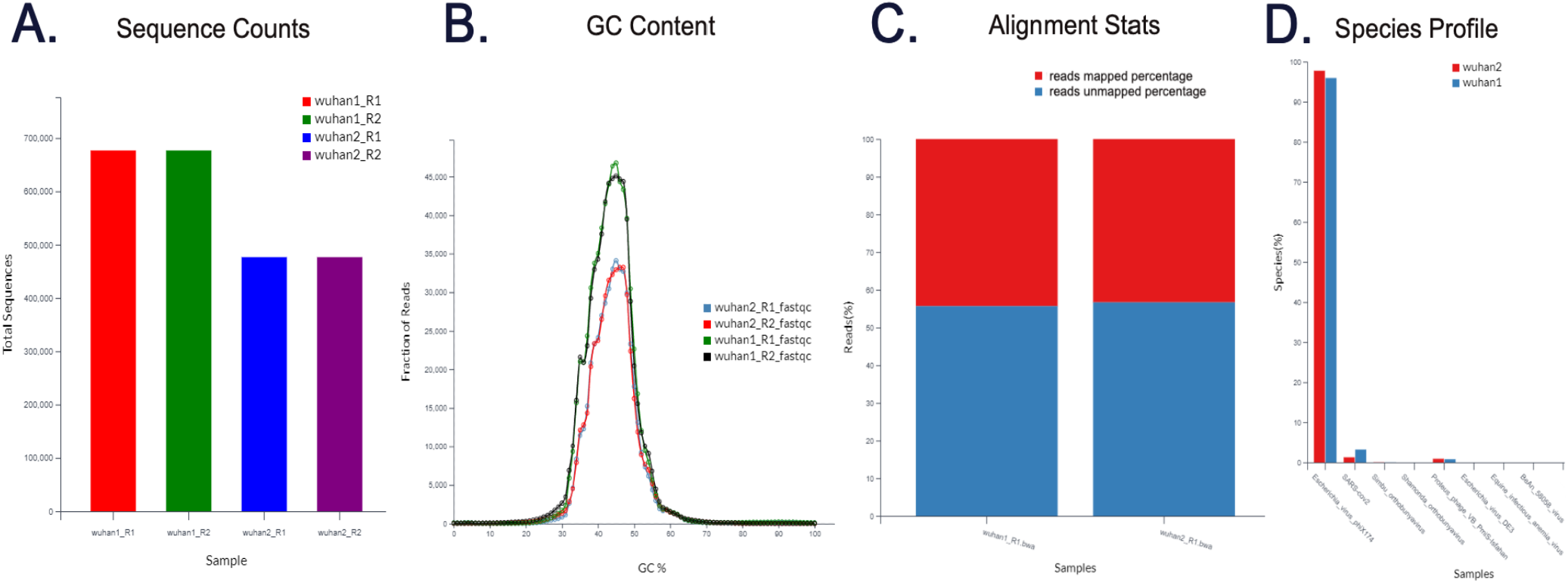
**A:** Sequence Counts of the patient samples. This plot will give a picture of how much data was generated by the sequencing machine for each sample in terms of number of reads. **B:** This plot shows the GC content across the whole length of each sequence. The GC content of the plot indicates the average GC percentage within each sample. The distribution should ideally show normal distribution. Any skew in the curve indicates either contamination or some problem with library preparation or PCR steps in the experiment. **C:** Alignment Stats. There are two levels of alignment, first, alignment to the human reference genome, and second the unmatched reads aligned to the coronavirus reference genome. The mapped sequences are taken for variant calling. **D:** Species Profile. The plot displays the pathogens identified in the patient sample (e.g. saliva swab); in this case both individuals sequenced have SARS-CoV2 virus sequences identified.

By pressing on a project, the user can see more details about the status of the computation and parameters used. If required, it is possible to download a log file and to monitor the costs of computations.

In case a pipeline fails, e.g. in the case there are problems with the raw data or metadata sheet, all processes the pipeline went through are monitored in real time with additional information on the computation’s CPU load and memory. In this way it is easy to find the step where a pipeline failed, and the error can be addressed.

After the pipelines are finished, the user can monitor the results by clicking the ‘Results’ button. All the pipeline-specific plots are presented in an interactive view and can be exported to a PDF report that contains all the parameters and settings of the analysis for easy results sharing.

By interrogating the species profile, the presence of SARS-CoV2 can be confirmed and the presence of other pathogens that may add to observed phenotypes can be analyzed. The other preinstalled pipelines also work with patient sequencing files:

### COVID-19 Assembler

This is a bioinformatics pipeline for SARS-CoV2 sequence assembly, by aligning and merging fragments from a longer viral DNA sequence in order to reconstruct the original sequence. The raw data, represented by FASTQ files, a mixture of human and pathogen sequences, are taken as input and then checked for quality. After removal of the adapter sequences, the reads are mapped to the human reference genome (hg19; GRCh37.75). The unaligned, viral sequences are then taken for de novo assembly using the Spades program and evaluated using the Quast program. The assembled genome files are viewable in programs such as Bandage. The contigs are subjected to gene/ORF prediction and the resulting sequences are further annotated using PROKKA.

### COVID-19 Alignment

This pipeline produces variant files and an annotation table from the SARS-CoV2 viruses present in the COVID-19 patient. Unlike going for a de novo assembly, the user can directly align the reads to the available pathogen reference genome and identify the variants presented within the analyzed samples. The resulting variant calls can be used in downstream analyses to identify clues regarding how the virus has evolved and the distribution of variants within the virus strain. The raw data (FASTQ short reads) from the patient are taken as input and are checked for quality with FastqQC. The reads are then mapped to the hg19 human reference genome. The unaligned reads are then taken for alignment with the coronavirus (NC_05512) reference genome. The alignments are checked for duplicates and realigned using Picard. The variants calling tools (lofreq) are used on these realigned files to get the list of the variants in the patient’s viruses. The variants are annotated with snpEff.

### COVID-19 Variant Caller (Patient)

This pipeline is designed to produce VCF variant files from the COVID-19 patients sequencing reads. The resulting VCF files can be valuable in identifying the variants associated with the coronavirus infection and can be easily exported and used for downstream analyses. The pipeline performs a quality check on the reads and trims the adapters. These trimmed reads are then aligned to the human reference genome (hg19). The alignment files are checked for duplicates, realigned and variants are called. These variants are annotated using the snpEFF program.

Overall, the whole workflow, from data upload to running a COVID-19 related pipeline was designed to be as easy as possible, with only a few steps involved and no command line entry needed, so that researchers and healthcare professional can run analyses of patients sequences and share the files and results with the research community. Researchers with other Nextflow pipelines related to tackle the COVID-19 pandemic are highly encouraged to share them on the Shivom platform.

## Results and Discussion

The main goal of this precision-medicine platform is to provide the global research community with an online marketplace for RAPID data sharing, managing & analytics capabilities, guidelines, pilot data, and deliver AI insights that can better understand complex diseases, aging & longevity as well as global pandemics at the molecular and epidemiological level.

Among other use-cases, the provided platform can be used to rapidly study SARS-CoV-2, including analyses of the host response to COVID-19 disease, establish a multi-institutional collaborative data-hub for rapid response for current and future pandemics, characterizing potential co-infections, and identifying potential therapeutic targets for preclinical and clinical development. No patient data can be traced back to the data donor (patient or data custodian) because only algorithms can access the raw data. Researchers querying the data-hub have no access to the genetic data source, i.e. no raw data (e.g. no sequencing reads can be downloaded). Instead, researchers and data analysts can run predefined bioinformatics and AI algorithms on the data. Only summary statistics are provided (e.g. GWAS, PHEWAS, polygenic risk score analyses). In addition to genetics bioinformatics pipelines, specialized COVID-19 bioinformatics tools are available (e.g., assembly and mapping pipelines). Using these free tools, researchers can quickly see why some people develop severe illness while others do show only mild symptoms or no symptoms whatsoever. We might hypothesize that some people harbor protective gene variants, or their gene regulation reacts differently when the COVID-19 virus attacks the host.

Particularly during an early outbreak, virus-host genetic testing has value in identifying those people who are at high or low risk of serious consequences of coronavirus infection^35^. This information is of value for the development of new therapies, but also for considering how to stratify risk, and identify those who might require more protection from the virus due to their genetic variants. Identifying variants associated with increased susceptibility to infection, or with serious respiratory effects, relies on the availability of genetic data from the affected individuals. However, here COVID-19 research encounters challenges associated with the population distribution of genetic data, and the consequent privileging of specific groups in genetic analyses^35^. Most data sets used for genome-wide association studies are skewed toward Caucasian populations, who account for nearly 80% of individuals in GWAS catalogs^40^. In comparison, those from African and Asian ancestry groups are poorly represented. Even large-scale genetic analyses may consequently fail to identify informative variants whose frequency differs among populations, either under- or overestimating risk in understudied ethnic subgroups.

Data from many countries showed that people from black and minority ethnic (BAME) groups have been disproportionately affected. People from a BAME background make up about 13% of the UK population but account for a third of virus patients admitted to hospital critical care units. Black Americans represent around 14% of the US population but 30% of those who have contracted the virus. More than 70% of healthcare professionals who have died in the UK have been from BAME backgrounds. Similar patterns showing disproportionate numbers of BAME virus victims have emerged in the US and other European countries with large minority populations. In the US and UK, the proportion of critically ill black patients is double that of Asians, so geography and socioeconomic factors alone are unlikely to explain the stark differences.

The key to fighting early disease outbreaks is speed. With the reality that some infections may originate from unknowing carriers, FAST patient stratification and identification is even more paramount. It is important to investigate if the severity of the disease may lie in the interaction with the host genome and epigenome. Why do some people get severely sick and die while others do not show any symptoms? It could be that some people harbor protective gene variants, or their gene regulation is able to deal with viruses attacking the host. Very quick data sharing and analysis ensures that precision medicine is brought to patients and healthy individuals faster, cheaper, and with significantly less severe adverse effects, leveraging information from the interaction between labs, biobanks, clinics, CROs, investigators, patients and a variety of other stakeholders.

### Changing the global data-sharing paradigm

As outlined in this paper, we have the technology to make easy data sharing a reality. However, the observation that data sharing technologies are still extremely underutilized and most data custodians are reluctant to share their data, demonstrates that the global healthcare community needs an elemental paradigm shift, a major change in the concepts and practices of how data is shared and utilized, particularly during a pandemic. A paradigm shift, as outlined by the American physicist and philosopher Thomas Kuhn, requires a fundamental change in the basic concepts and experimental practices of a scientific discipline. Kuhn presented his notion of a paradigm shift in his influential book The Structure of Scientific Revolutions. We argue that the world needs such a revolutionary paradigm. Researchers use real-world data from laboratories, testing centers, hospitals, academic institutions, payors, wearables, and other data sources such as direct to consumer genetic companies to better understand disease outbreaks and to develop vaccines and medicines quickly. Such data is used across the whole spectrum of drug discovery, vaccine development, and basic academic research. However, whoever has ever applied for data access to a major biobank knows that typically, it’s costly, time-consuming and nerve wrecking, in the best-case scenario, to obtain such data.

Data is often received from a third-party partner that aggregates, anonymizes it, and makes it usable for their research purposes. Often, there is a significant lag time between when the data is generated and when it’s available for use. For example, when an infected patient is sequenced in a hospital, it can take between weeks and even years until the data surfaces and is available in a public database. But if healthcare systems are trying to control a pathogen outbreak and want to understand more about the affected patients and host-virus interactions, things are changing day by day, we need that insight into the real-time aspect of what’s going on.

What we need is an ecosystem that, for example, allows a doctor who sees a patient and sends a buccal swab out for sequencing, to get sequencing results, sharing it with the global community and gets in-depth data analytics back to evaluate if a similar host-virus interaction was observed anywhere else on the planet - within 24 hours or less. In the current pandemic, for example, there is a need to understand COVID-19 patient data at the local level, directly from the sources, to get data in real time. Omics data from patients, exposed individuals, and healthy control subjects will be a critical tool in all aspects of the response—from helping to design clinical trials, identifying vulnerable groups of the population, to understanding whether COVID-19 patients taking specific medicines are at higher risk of developing severe side-effects.

### Incentives for sharing data

The ability to collect, analyze, and interpret data is fundamental to the management of infectious diseases. To do so, stakeholders need to be incentivized to share their data. Sharing data is good for science, not only because transparency enhances trust in science, but because data can be reused, reanalyzed, repurposed, and mined for further insight. Sharing data on the Shivom platform can increase transparency. The more transparent the contributors are about their data, analytic methods, or algorithms, the more confident they are of their work, and more open to public scrutiny. Using the platform can also help researchers and consortia that want to collaborate within a safe space. We need more data aggregators that are committed to supporting an ecosystem of research communities, businesses, and research partners, by sharing data or algorithms in safe and responsible ways. Such an open ecosystem approach can yield high dividends for society. In addition, consortia can come together to build patient cohorts, maximizing their research budget. Research organizations who share similar goals can join efforts to create insight at scale, creating datasets that meet the needs of multiple stakeholders at lower costs.

A successful response to the COVID-19 pandemic requires convincing large numbers of scientists and healthcare organizations to change their data sharing behaviors. Although the majority of countries have population studies and build massive digital biobanks, to our knowledge, none of the larger biobanks in the world have a data sharing infrastructure to share data with other countries. The Shivom platform provides the only cross-border data marketplace to allow data custodians and biobanks to easily & securely share and monetize anonymized datasets. A massive, decentralized, and crowd-sourced data can reliably be converted to life-saving knowledge if accompanied by expertise, transparency, rigor, and collaboration. Overall, there are several obvious advantages using the platform:

- **Making data actionable** as data can be analyzed on the bioinformatics platform
- **Breaking down data silos**, putting data in a larger, global context
- Making data easily **findable & searchable**
- Ability to easily **share data with fine-grained permission** with clients, collaborators, or the public
- **Improved accessibility** by avoiding complicated access procedures, promoting maximum use of research data
- **Advanced interoperability** by integrating with applications or workflows for analysis, storage, and processing; e.g. new AI algorithms
- **Better reusability** so that data is not stored and forgotten, but reusable in many research projects
- **Increased impact** as sharing data will help the global fight against complex and rare diseases
- **Gain reputation** for the user’s organization by giving researchers the ability to find, use and cite their work

### Data Monetization Options

Usually, for most data custodians such as clinical laboratories, data clean-up procedures and data deposition are not formal responsibilities. These jobs have to be done by some fairly busy people, in an increasingly demanding environment. There is no remuneration for the labs to submit their data; but done out of goodwill. As long as the data collection is funded by public institutions this is no major problem, but private hospitals, small biotech or genetic companies, data sharing can become a challenge. The Shivom platform offers the possibility to monetize their datasets. Practically, that means if a dataset is monetizable, whenever a bioinformatics pipeline or algorithm touches the dataset, a smart contract is triggered that sends a predefined fee to the data custodian. The pricing can be adjusted depending on the quality, data type and other factors.

While open data sharing is encouraged for the publicly funded research community, such monetization options can be a valuable extra resource of funds for organizations that have to worry about sustainability and revenue models. Unlike many other data brokers, organizations can upload and monetize data directly without a distributor or middleman. This leads to real-time payouts, higher revenue share rates, and instant upload availability. It allows researchers to upload their own data and get it in front of an audience of other scientists. In a way, it almost acts like a content creation tool, rather than a pure data sharing service.

From the data-user perspective (e.g. a pharma or biotech company), getting access to large genetic datasets gets much easier and more affordable as it is not necessary to acquire exclusive licenses that can cost millions of dollars. Instead, the data user only has to pay for accessing the data that is actually used, similar to the way people access music on streaming services. In addition, since the data custodian remains full ownership of the data, there is no need for complicated business relationships, nor will their internal business model be challenged.

### Tackling Data Curation Dept

Using the Shivom platform as the regular data repository can help researchers with their data curation tasks, preventing what was termed ‘data curation dept’^41^. Researchers can migrate/copy their data to the platform to protect themselves from breaking in-house legacy hardware, e.g. old unmaintained servers. Also, digitizing raw data and storing it increases security of the data as many datasets are still stored on decaying physical media (e.g. consents done on paper, metadata stored on CDs/DVDs, MiniDisc, memory sticks, external hard drives etc.). If the physical medium decays, then the raw data are lost, making it impossible to reproduce the original research or apply new techniques to analyze the original raw data.

Loss of data has far-reaching consequences: a recent survey demonstrated that the lack of availability of raw data and data provenance were common factors driving irreproducible research^42^. For many academic labs, the loss of data could lead to a loss of funding because future grant applications would not be able to list their data as a resource. Even for normal daily tasks in a typical university laboratory, if data is not lost but requires additional work to locate, for example, after a PhD student left the research group, then an additional time and staff cost is involved to find the data and re-learning knowledge that was the domain of the departed team member. Particularly in multinational pandemic cohort studies where the recruitment and data collection are so difficult, it is unacceptable that there is such a risk to the hugely valuable resource of historical data through the accrual of data curation debt^41^.

Joining forces and sharing information at the national level also eases and supports international cooperation initiatives. National coordination of pandemic policy responses can also benefit from joining forces with international research cooperation platforms and initiatives. It is important to note that data stored and shared on the Shivom platform does not need to be exclusive to this data marketplace; the data can be hosted in other databases as well.

The benefits of data sharing do not end here, other benefits that should incentivize researchers to share their datasets include the chance of getting more citations, increase of exposure that may lead to new collaborations, boosting the number of publications, empowering replication, avoiding duplication of experiments, increasing public faith in science, guiding government policy, and many more. Also, far from being mere rehashes of old datasets, more and more evidence shows that studies based on analyses of previously published data can achieve just as much impact as original projects and published papers based on shared data are just as likely to appear in high-impact journals, and are just as well-cited, compared with papers presenting original data^43,44^.

Building a massive global data-hub with its wealth of multi-omic information is a powerful tool to study, predict, diagnose and guide the treatment of complex disease and help us to live longer and healthier. Also, data sharing should not stop with a successful vaccine rollout.

Healthcare systems need to monitor closely the characteristics of the most promising vaccine candidates. Such monitoring includes understanding the patient’s response, dosing regimen, potential efficacy, and side effects. Measures to monitor and control the COVID-19 pandemic by the scientific community will be relevant for as long as its risk continues.

### Regulatory and ethical consideration

Moving genomics data and other information into routine healthcare management will be critical for integrating precision medicine into health systems. However, data sharing is often hindered by complicated regulatory and ethics frameworks. Obviously, there are ethical and legal issues to consider in any work that involves sharing of sensitive personal data. Nevertheless, it is obvious that we need to be able to share data to fight global pandemics. The SARS-CoV2 virus does not care about jurisdictional boundaries. Data from COVID-19 patients enables experts to interpret complex pieces of biological evidence, leading to scientific and medical advances that ultimately improve care for all patients. But these advantages must be balanced against the duty to protect the confidentiality of each individual patient.

It is out of the scope of this study to discuss ethics in depth. The Shivom platform is built to be operational in all jurisdictions around the world. Many regulatory frameworks do already exist that allow to collect and hold anonymized data from patients in the interest of improving patient care or in the public interest. It is now time to go a step further and decouple procedures from regional regulatory frameworks, to make data sharing easy across borders and regulatory frameworks. In this context, the magic word is anonymization.

On the outlined data sharing platform, no access to raw data is permitted. Only algorithms that provide summary statistics are able to touch the data, guaranteeing that no dataset can be traced back to an individual. This process makes it possible to share important multi-omics information anonymously and securely, whilst still enabling linkage to metadata such as diagnosis, lifestyle factors, treatment and outcome data. The key is that data on the platform is largely protected from re-identification of individuals. Re-identification in this context is understood as either identity disclosure or attribute disclosure, that is, as either revealing the identity of a person, thus breaching his or her anonymity, or disclosing personal information, such as susceptibility to a disease, which is a breach of privacy^20^. The Shivom platform was designed to make it impossible to disclose information from genetic data not solely on the individual, but also on their relatives and their ethnic heritage, e.g. protecting the identity of siblings or other relatives in the context of forensic investigations.

Ideally, patients should be involved in such data sharing initiatives. Institutions that collect and host data and promote data sharing should explain the benefits of collecting and using patient data to their clients. There is a need for public education on genetics, to take the frightening aspect away, ultimately incentivizing patients and healthy control groups to share their data anonymously for the greater good.

### Easy Public Access

One of the cornerstones of the proposed data ecosystem is easy public access to data. If adopted by the research community, researchers seeking to use public data must no longer comb websites and public repositories to find what they need. It is important that every human being has the potential to participate in this process, either on the data provider side or by analyzing datasets. Access to the data should not be restricted to certain organizations or researchers with a long publication record. These systems are outdated and archaic.

Usually, with most databases and biobanks, the bona fide user must apply for access with a corresponding data access control body, provide an institution ID such as OpenID or ORCID, and often provide a long publication record. Usually, that means they must register from, and be affiliated with, an approved research institute and need to register their organization which means they will need their organization’s signing official to participate in the registration process. Often, when interested researchers want to access data in public databases, they are asked to submit a lengthy proposal detailing the data they want to use and the rationale for their request. Most of the time, projects must involve health-related research that is in the public interest. Sometimes the researcher needs to go through an Institutional Review Board (IRB) first and even then, all proposals received by the data repository are considered on an individual basis and may have to be evaluated by an internal or external ethics committee. On average, a straightforward application and approval process takes 2–3 months; sometimes it has taken up to 6 months^45^. Obtaining access to data that are available from international sources adds another layer of complexity, due to international law.

Most databases require the user then require a standard material transfer agreement (MTA) to be signed prior to any data delivery that governs how the researcher can use the data. But even when data access is granted, it is common that datasets are restricted to only those data and participants that the researcher required at that time (e.g. COVID-19 patients only or specific case–control subsets). In addition, most agreements grant access for a limited time only, at the end of which a report summarizing research progress is required; for continued data access, yearly renewal requests are often obligatory.

To exemplify the problem, our researchers applied for access to COVID-19 data with four data-providers (e.g. data repositories, COVID-19 research consortia, and biobanks) in the UK, EU and US to support our COVID-19 research efforts. All four requests were rejected, each for a different reason. One public data repository denied access to their data because their legal terms only allow access for groups working in public institutions but not private/commercial companies. Another biobank was concerned about the lack of publications from our research unit that could demonstrate the organization’s current activity in health research, keeping in mind that our researchers are highly respected in their field and have a vast list of scientific publications from previous appointments. Other objections were that the proposed COVID-19 research was too broad, basically no exploratory studies were permitted, or that our junior researcher had no publication record (5 or more papers). One COVID-19 data consortium suggested that our organization shares our genetic data with them but did not allow access to their data sets and only provided access to secondary analyses that their researchers performed on the data. Such processes are often undemocratic, against the public interest and simply take too long to provide any benefit under a health crisis.

A large majority of biobanks and databases are non-profit organizations, and most of them operate on a not-for-profit basis. Consequently, public funding puts some obligation on data-custodians to enable research with any scientific and social value. Against this background, we argue requiring data custodians to prioritize access options and avoid underutilization of data. Thus, biobanks ought to make all necessary arrangements that facilitate the best possible utilization of the data sets.

As such, interested researchers on our data hub do not require a long publication record, nor do they need to provide a 30-page proposal, referrals, or a membership to an elite club (e.g. an accredited, approved research institute) to search and analyze data. Usability and accessibility needs to be guaranteed for every interested individual, the academic researcher from any elite university as well as the pharma manager, the PhD student from Kenia, the doctor at a local clinic in Wuhan, a student in a remote area of Pakistan, or the nurse in a village hospital in Brazil. In addition, data should not only be accessible for a specific research purpose but should enable research that is broader in scope and exploratory in nature (i.e. hypothesis-generating).

### Other Use cases

Many diseases hide in plain sight – in our genome – making genomics the most specific and sensitive way to identify disease early and guide both patients and healthcare practitioners to the best course of treatment. Yet despite many advances in data analytics and AI, our understanding of the human panome (including epigenome, proteome, metabolome, transcriptome, and so forth) is incomplete and the ways in which we utilize this information is limited and imperfect.

#### Clinical Diagnosis

Having a secure data-hub for fast data dissemination and analytics in the clinical setting will improve routine clinical care. Most complex diseases, such as cancer have become increasingly dependent on genetic and genomic information. When biopsy samples are collected from patients and sequenced, there is the need to be able, as quickly as possible, to tell whether any variants detected are associated with the disease or indicate that a patient is likely to respond to a particular drug. At the same time, with every sequence shared, the accuracy of the global diagnostic accuracy improves, not only for the current patient but for all cancer patients around the globe. In addition, linking patients’ clinical data with genomic data will help answer research questions relevant to patient health that would otherwise be difficult to tackle.

#### Rare Diseases

Access to genetic data and ‘real-world evidence’ (RWE) obtained from observational data is crucial to help push research and innovation particularly on rare diseases. The standard approach to clinical trials is challenging, if not impossible for most rare diseases. It is the characteristics of rare diseases that they can only be adequately tackled if there is enough data to make evidence-based decisions, given that the number of patients is so small. To make this happen it is absolutely necessary to break down global data silos. To move things forward, many global initiatives, such as the EU Regulation on Orphan Medicinal Products (OMPs) were introduced specifically to address the challenge of developing medicines that treat patients with rare diseases. However, the initial progress seen since the introduction of these initiatives has tapered off in recent years, with a decline in a number of drug approvals over the past years. In fact, 95% of rare diseases conditions remain without treatment.

One of the main hurdles is the collection and access to data, particularly obtained outside the context of randomized controlled trials and generated during routine clinical practice. Gaining the critical mass of data to close data gaps needs global coordination and represents a critical step for driving forward research in the area of orphan drugs.

In addition, the described platform can be used to collect valuable datasets to specific themes and therapeutic areas that are not yet available. Currently, several pilot data initiatives started to collect research-area specific datasets on the Shivom platform:

#### Cannabis Data-Hub

In collaboration with direct to consumer companies, patient support groups and medical professionals, we aim at collecting data on the interaction of individual’s genomes and medicinal compounds found in medical and recreational cannabis. Cannabis-related research allows governments and pharma researchers to better understand efficacy and potential side effects of hundreds of compounds like cannabinoids and terpenes and to improve legal frameworks for cannabis products. For example, several gene variations are known to affect the user’s endocannabinoid system, increasing sensitivity to Δ9-Tetrahydrocannabinol (THC), potentially posing risk factors for THC-induced psychosis and schizophrenia^46,47^. Similarly, certain variations in AKT1 can make people’s brains function differently when consuming marijuana^48^. About 10 percent of occasional cannabis users develop a physiologic dependence, cravings, or other addiction-related behaviors that can affect everyday life. As a result, many cannabis users often go through various products blindly before their ideal balance between their drug’s efficacy and their tolerability is reached. Patients and their doctors can learn about pharmacogenetic aspects of cannabis use and be informed on their susceptibilities and can therefore adjust cannabis habits to best fit their genotype. Having a comprehensive anonymized genetic database available combined with real world data will pave to way to better pain management as it’s been demonstrated that cannabis may be a promising option to replace opiates in the treatment of pain, as well as replacing other drugs such as antiepileptics and antipsychotics^49^.

#### The Longevity Dataset

is an ongoing, concerted effort to provide genetic and epigenetic information on individuals who have attained extreme ages. The remarkable growth in the number of centenarians (aged ≥ 100) has garnered significant attention over the past years. Consequently, a number of centenarian studies have emerged, ranging in emphasis from demographic to genetic. However, most of these studies are not aimed at breaking down data silos. As such, data will be collected from centenarians, with a subset of people who attained an age of 110 years or higher – so-called “supercentenarians” to yield sufficient numbers to warrant descriptive studies and to find genetic factors associated with exceptional longevity.

Having enough datasets will improve the search for meaningful subnetworks of the overall omic network that play important roles in the supercentenarians’ longevity^50^ and will help to better understand epigenetic mechanisms of aging^51,52^.

#### A data repository for the scientific publishing process

Having data deposited in a public data-hub is of utmost importance, particularly when data needs to be published rapidly, as in a time sensitive pandemic. The case is easily demonstrated by retraction of two COVID-19 studies in May of 2020^53^. The New England Journal of Medicine published an article that found that angiotensin converting enzyme (ACE) inhibitors and angiotensin receptor blockers were not associated with a higher risk of harm in patients with COVID-19^54^. The Lancet published an observational study by the same authors, indicating that hospital patients with COVID-19 treated with hydroxychloroquine / chloroquine were at greater risk of dying and of ventricular arrhythmia than patients not given the drugs^55^. Both journals retracted these articles because the authors said they could no longer vouch for the veracity of the primary data sources, raising serious questions about data transparency. The problem was that both studies used data from a healthcare analytics company called Surgisphere. After several concerns were raised with respect to the correctness of the data, the study authors announced an independent third-party peer review. However, the data custodian Surgisphere refused to transfer the full dataset, claiming it would violate confidentiality requirements and agreements with clients, leading the authors to request the retraction of both studies. This problem could have been avoided by publishing the datasets to a data repository as the described platform in this article. Journal reviewers and other researchers could have easily accessed and analyzed the data before problems arose, a method that could easily be a mandatory part of the submission process.

The urgency of sharing emerging research and data during a pandemic is obvious. Rapid publication and early sharing of results is clearly warranted, but it means that the research community must also be able to access and analyze the data in a timely manner. Publishing research data to an open platform with audit features and data provenance increases the chance of reproducibility and to have reliable assurances for the integrity of the raw data. Considering that we have the technical capabilities to publish anonymized datasets, journals, in principle, should try to have their authors publicize raw data in a public database or journal site upon the publication of the paper to increase reproducibility of the published results and to increase public trust in science^56^. In this context, sharing raw omics data publicly should be a necessary condition for any master or PhD thesis as well as research studies to be considered as scientifically sound, unless the authors have acceptable reasons not to do so (e.g. data contains confidential personal information); keeping in mind that fine-grained permission settings also allow to publish and analyze industrial & proprietary research.

Overall, by utilizing an easy-accessible data-hub, it is possible to incorporate valuable omics information into routine preventive care, research and longitudinal patient monitoring. In the end, we can leverage the benefits of omics to expand patient access to high-quality care and diagnosis.

### Next Steps

It is the aim of the authors to further evolve the proposed data sharing and analytics platform to the point that it is a widely used and truly actionable data hub for the global healthcare ecosystem. Together with the research community, we are working on adding a variety of other functionalities to make this precision medicine ecosystem more valuable and innovative, particularly in the fields of artificial intelligence, interoperability, pandemic preparedness, and other new research fields. Long-term this can include projects to add functionalities for clinical trials, AI marketplaces, IoT data, or quantum computing. Some of the planned addons in the roadmap for future releases include a cohort browser, improved search engines, many new pipelines and AI tools, as well as fully federated data sharing capability. We plan to add a forum to the platform to help researchers exchange information on datasets and data curation. Often, those who want to make their data more open face a bewildering array of options on where and how to share it. Providing a space for sharing data and guiding young researchers in their task by sharing necessary expertise in data curation and metadata will help to ensure that the data they plan to share are useful for others.

### COVID-19 Call to action

The urgency of tackling COVID-19 has led governments in many countries to launch a number of short-notice and fast-tracked initiatives (e.g. calls for research proposals). Without proper coordination amongst ministries and agencies, they run the risk of duplicating efforts or missing opportunities, resulting in slower progress and research inefficiencies. The best way to prevent a further spread of the virus and potential new pandemics is to apply the same principles as we use to prevent catastrophic forest fires: survey aggressively for smaller brush fires and stomp them out immediately. We need to invest now in an integrated data surveillance architecture designed to identify new outbreaks at the very early stages and rapidly invoke highly targeted containment responses. Currently, our healthcare system is not set up for this. First, modern molecular diagnostic technologies detect only those infectious agents we already know exist; they come up blank when presented with a novel agent. Infections caused by new or unexpected pathogens are not identified until there are too many unexplained infections to ignore, which triggers hospitals to send samples to a public health lab that has more sophisticated capabilities. And even when previously unknown infectious agents are finally identified, it takes far too long to develop, validate and distribute tests for the new agent. By then, the window of time to contain an emerging infection will likely be past. Using the platform as demonstrated in this article, all those processes can be sped up dramatically. It would enable our healthcare ecosystem to halt the spread of emerging infectious diseases before they reach the pandemic stage. In this new surveillance architecture proposed here, genomic analysis of patients presenting with severe clinical symptoms would be performed up front in hospital labs piggybacked on routine diagnostic testing.

Researchers can use the data in our data-hub to compare new cases with existing datasets. Given national epidemiological data on infection rates; where symptomatic people seek healthcare, a remarkably small number of surveillance sites would be required in order to identify an outbreak of an emerging agent. Using genomic sequencing, the novel causative agent could be identified within hours, and the network would instantaneously connect the patients presenting at multiple locations as part of the same incident, providing situational awareness of the scope and scale of the event.

Using a globally accessible, easy-to-use solution that provides a ‘one-stop shop’ for the centralization of information on new virus-host interactions can help ensure that appropriate conditions for collaborative research and sharing of preliminary research findings and data are in place to reap their full benefits. Beyond short-term policy responses to COVID-19, incorporating data sharing with the Shivom platform into ongoing mission-oriented research and innovation policies could help tackle future pandemics on a national or international scale, for example to enhance datasets informativeness to reveal host infection dynamics for SARS-CoV-2 and therefore to improve medical treatment options for COVID-19 patients. In addition, it can help to determine and design optimal targets for therapeutics against SARS-CoV-2.

The platform is in alignment with recent guidelines published by the Research Data Alliance (RDA) COVID-19 Working Group^57^. The RDA brought together various, global expertise to develop a body of work that comprises how data from multiple disciplines inform response to a pandemic combined with guidelines and recommendations on data sharing under the present COVID-19 circumstances. The RDA report offers best practices and advice – around data sharing, software, data governance, and legal and ethical considerations – for four key research areas: clinical data, omics practices, epidemiology and social sciences. In line with these recommendations, the Shivom platform can be used for:

- Sharing clinical data in a timely and trustworthy manner to maximize the impact of healthcare measures and clinical research during the emergency response
- Encouraging people to publish their omics data alongside a paper
- Underlining that epidemiology data underpin early response strategies and public health measures
- Providing a platform to collect important social and behavioral data (as metadata) in all pandemic studies
- Evidencing the importance of public data sharing capabilities alongside data analytics
- Offering a general ecosystem to exploit relevant ethical frameworks relating to the collection, analysis and sharing of data in similar emergency situations

At this point in time, the research community should be clear that a second or third wave of the COVID-19 pandemic will hit most countries. In addition, other epidemics and global pandemics will follow; it is just a matter of time. As such, the global research and medical community need to be prepared. When analysis of datasets shared on the Shivom platform results in publications, we encourage the researchers to cite this publication to enhance widespread adoption of this data sharing and analytics marketplace. We also encourage sharing data from COVID-19 and other virus outbreaks, including data from people who were exposed to the same pathogen but did not get the disease or possibly were infected but did not get symptoms.

## Conclusion

We developed a next-generation data sharing- & data analytics ecosystem that allows users to easily and rapidly share and analyze omics data in a safe environment and demonstrate its usefulness as a pandemic preparedness platform for the global research community. Adopting the platform as a data hub has the clear benefit that it enables individual researchers and healthcare organizations to build data cohorts, making large, or expensive-to-collect, datasets available to all. In this way, data sharing opens new avenues of research and insights into complex disease and infectious disease outbreaks. This is not just true of large-scale data sharing initiatives, even relatively small datasets, for example rare disease patient data, if shared, can contribute to big data and fuel scientific discoveries in unexpected ways. Particularly during early disease outbreaks, the patient-level meta-analysis of similar, past outbreaks may reveal numerous novel findings that go well beyond the original purpose of the projects that generated the data. Using the platform as demonstrated will enhance opportunities for co-operation and exploitation of results. Our data sharing approach is inclusive, decentralized, and transparent and provides ease of access. If adopted, we expect the platform to substantially contribute to the understanding of the variability of pathogen susceptibility, severity, and outcomes in the population and help to prepare for future outbreaks. In providing clinicians and researchers with a platform to share COVID-19 related data, the clinical research community has an improved way to share data and knowledge, removing a major hurdle in the rapid response to a new disease outbreak.

## Supporting information

Metadata structure

Metadata example file

Metadata template

## Data Availability

No external datasets.

## Author Contributions

Conceived and designed the platform: AS, IF. Analyzed data: IF, GL, DB. Wrote the paper: AS.

