## Supplementary material for "A global omics data sharing and analytics marketplace: Case study of a rapid data COVID-19 pandemic response platform": Metadata structure

### Supplement

**S1. COVID19 Metadata Sheet. V1.0**

These values should be collected in an ideal case scenario for describing COVID19-exposed individuals. Please note that not all values are necessary. The metadata-template can be downloaded here (xls/csv). When a .vcf from a COVID-19 patient is uploaded, the user should select: "COVID-19" under "Health Status" in the import record dialogue. If available, we recommend adding more metadata from the list collected by the Genetics Host Initiative. The metadata sheet can be found here.

| Metadata field | Potential Values |
|---|---|
| `filename` | Example: `patient057.vcf` or `345_23andme65776.vcf`<br>This entry is linked to the vcf file in the database or to the file that was uploaded. |
| `height` | The current height of the individual in centimetres `[cm]` |
| `weight` | The current weight of the individual in kilograms `[kg]` |
| `num_dependents` | Number of dependents in the family. A dependent is a person who relies on another as a primary source of income. For example, minors (children who are under the age of majority) are dependants of their parents or legal guardians.<br>Value: `[Number]` |
| `religion` | The current religion of the individual.<br>Values:<br>`Christianity, Islam, Secular, Nonreligious, Agnostic, Atheist, Hinduism, Buddhism, Chinese traditional religion, Ethnic religion, Sikhism, African traditional religion, Spiritism, Judaism, Bahá'í, Jainism, Shinto, Cao Dai, Zoroastrianism, Tenrikyo, Neo-Paganism, Unitarian Universalism, Rastafari` |
| `ethnicity` | The self-reported ethnicity of the individual.<br>Values:<br>`East Asian, South Asian, White/Caucasian, Mixed (Any combination of backgrounds), Hispanic/Latino, Black, Middle Eastern or North African, Other, Prefer not to say` |
| `family_status` | The marital status of the individual.<br>Values:<br>`Married, Common Law, Single, Never Married, Widowed, Divorced, Separated` |
| `is_smoker` | The lifestyle factor smoking of the individual.<br>Values:<br>• `Current, everyday smoker`<br>• `Current, some day smoker`<br>• `Former smoker`<br>• `Never smoker`<br>• `Smoker, current status unknown`<br>• `Unknown if ever smoked` |
| `is_drinker` | The lifestyle factor drinking of the individual.<br>Values:<br>• `Current, drink every day`<br>• `Current, drink some day`<br>• `Former drinker`<br>• `Never drinker`<br>• `Drinker, current status unknown`<br>• `Unknown if ever drank` |
| `is_drug_user` | The lifestyle factor drinking of the individual.<br>Values:<br>• `Current, everyday drug user,` |

| | |
|---|---|
| | • Current, occasional drug user,<br>• Former drug user,<br>• Never used recreational drugs |
| education_degree | The formal education of the individual if known (highest education level).<br>Values:<br>• Grade 10 (high school sophomore) or less<br>• Grade 11 or 12 (high school junior or senior)<br>• Vocational/technical school/some college or university<br>• Bachelor's degree or higher |
| excercise | The usual physical activity of the individual.<br>Values:<br>• Poor - no regular physical activity<br>• Moderate - some physical activity<br>• High - regular physical activity |
| dietary_regimen | The current diet of the individual. Values:<br>• Lactose-free,<br>• Low carb,<br>• Low fat,<br>• No Special diet,<br>• Paleo,<br>• Vegan,<br>• Vegetarian |
| gender | The gender (sex) of the individual.<br>Values:<br>• Male<br>• Female<br>• Transgender<br>• Other |
| comorbidity1 | Diagnosis of main comorbidity (disease). If possible, using standards, i.e. Human Disease Ontology/DOID |
| comorbidity2 | Diagnosis of a second comorbidity (disease). If possible, using standards, i.e. Human Disease Ontology/DOID |
| comorbidity3 | Diagnosis of a third comorbidity (disease). If possible, using standards, i.e. Human Disease Ontology/DOID |
| chcmi | The **Charlson Comorbidity Index** predicts 10-year survival in patients with multiple comorbidities. https://www.mdcalc.com/charlson-comorbidity-index-cci<br>Value: [%] |
| pneumst | What is (was) the **Pneumonia Status** of the individual?<br>• Non-present<br>• Present unilateral<br>• Present bilateral (dual) |
| sofa | The Sequential Organ Failure Assessment (SOFA) Score is a mortality prediction score that is based on the degree of dysfunction of six organ systems. The score is calculated on admission and every 24 hours until discharge using the worst parameters measured during the prior 24 hours. https://www.mdcalc.com/sequential-organ-failure-assessment-sofa-score<br>Value: [SOFA score (points)] |
| cpis | The **Clinical Pulmonary Infection Score (CPIS)** was developed to serve as a surrogate tool to facilitate the diagnosis of ventilator-associated pneumonia (VAP). The CPIS is calculated on the basis of points assigned for various signs and symptoms of pneumonia (eg, fever and extent of oxygenation impairment). https://www.mdcalc.com/clinical-pulmonary-infection-score-cpis-ventilator-associated-pneumonia-vap<br>Value: [CPIS score (points)] |
| qsofa | The quick Sepsis Related Organ Failure Assessment (qSOFA) score is a bedside tool to identify patients with suspected infection outside the intensive care unit who are at |

| | |
|---|---|
| | greater risk for a poor outcome. The qSOFA score can be used to identify patients at high risk of death. https://qsofa.org/<br>Value: **[qSOFA score (points)]** |
| cpap | **Continuous positive airway pressure (CPAP)** applied<br>• **Yes**<br>• **No**<br>• **Unknown** |
| antiviral | **Antiviral at diagnosis** using the Drug Ontology (DRON) https://www.ebi.ac.uk/ols/ontologies/dron<br>Value: **[Name]** |
| dayssympt | Days with symptoms of the individual<br>Value: **[Number of days]** |
| Mortality1 | Did the individual die from the condition?<br>• **Yes**<br>• **No** |
| twin | Is the individual a one- or two-egg twin.<br>Values:<br>• **No, not a twin**<br>• **Yes, One-egg twin**<br>• **Yes, Two-egg twin**<br>• **Yes, I don't know, either one-or-two** |
| handedness | Is the individual by birth left-, right- or both-handed?<br>• **Left-handed**<br>• **Right-handed**<br>• **Both-handed**<br>• **Unknown** |
| exposed_carrier | Exposed to known COVID19 carrier in the last 20 days.<br>• **No**<br>• **Yes**<br>• **Unknown** |
| travel | Travel (in the last 20 days)<br>• **Domestic**<br>• **International**<br>• **Domestic and International**<br>• **None**<br>• **Don't know** |
| job_medical | Is the individual working as medical professional?<br>• **No**<br>• **Yes**<br>• **Prefer not to answer**<br>• **Don't know** |
